# Sensitivity to missing not at random dropout in clinical trials: use and interpretation of the Trimmed Means Estimator

**DOI:** 10.1101/2021.03.05.21252334

**Authors:** Audinga-Dea Hazewinkel, Jack Bowden, Kaitlin H. Wade, Tom Palmer, Nicola Wiles, Kate Tilling

**Affiliations:** Population Health Sciences, Bristol Medical School, University of Bristol, Bristol, U.K.; Medical Research Council Integrative Epidemiology Unit, Bristol Medical School, University of Bristol, Bristol, U.K.; Centre for Academic Mental Health, Population Health Sciences, Bristol Medical School, University of Bristol, Bristol, U.K.; Exeter Diabetes Group (ExCEED), College of Medicine and Health, University of Exeter, Exeter,U.K.

**Keywords:** Trimmed means, dropout, randomized controlled trials, missing not at random, sensitivity analyses, bias quantification

## Abstract

Outcome values in randomized controlled trials (RCTs) may be missing not at random (MNAR), if patients with extreme outcome values are more likely to drop out (e.g., due to perceived ineffectiveness of treatment, or adverse effects). In such scenarios, estimates from complete case analysis (CCA) and multiple imputation (MI) will be biased. The trimmed means (TM) estimator operates by setting missing values to the most extreme value, and then “trimming” away equal fractions of both treatment groups, estimating the treatment effect using the remaining data. The TM estimator relies on two assumptions, which we term the “strong MNAR” and “location shift” assumptions. In this article, we derive formulae for the bias resulting from the violation of these assumptions for normally distributed outcomes. We propose an adjusted estimator, which relaxes the location shift assumption and detail how our bias formulae can be used to establish the direction of bias of CCA, MI and TM estimates under a range of plausible data scenarios, to inform sensitivity analyses. The TM approach is illustrated with simulations and in a sensitivity analysis of the CoBalT RCT of cognitive behavioural therapy (CBT) in 469 individuals with 46 months follow-up. Results were consistent with a beneficial CBT treatment effect. The MI estimates are closer to the null than the CCA estimate, whereas the TM estimate was further from the null. We propose using the TM estimator as a sensitivity analysis for data where it is suspected that extreme outcome values are missing.

## 1 Introduction

Randomized controlled trials (RCT) are considered the gold standard for assessing causality, because the randomization process ensures that unmeasured characteristics are well-balanced across groups. However, RCTs remain vulnerable to other sources of bias, including those which arise from missing data due to dropout. We focus here on the case where an RCT has missing values solely in the outcome, for example due to drop out.

The impact of missing data depends on the missingness mechanism and the analysis model.^1^ Three distinct missingness mechanisms can be distinguished: missing completely at random (MCAR), missing at random (MAR) and missing not at random (MNAR). With MCAR, missingness is unrelated to any measured or unmeasured characteristics and the observed sample is a representative subset of the unobserved full data.^3^ MAR means the missingness can be explained by observed data and, with MNAR, missingness is a function of the unobserved data (in our case, the outcome) itself.^2^

There are three main ways of dealing with incomplete data: a complete case analysis (CCA), inverse probability weighting (IPW) and multiple imputation (MI). A CCA is the analysis model intended to be applied to the trial data at its outset, restricted only to individuals with observed outcomes. Assuming correct model specification, CCA is unbiased if the outcome is MCAR or MAR^3^. With IPW, the same analysis model is fitted to the complete cases, but each individual is now weighted by the inverse of its probability of being observed. With MI, the observed data are used to repeatedly predict - “impute” - the missing values. The analysis model is applied to multiple imputed datasets and the estimates pooled using Rubin’s rules.^1^. Broadly, IPW and MI will be valid if the data are MAR and the weighting and imputation models are correctly specified, respectively. When data are MNAR, however, the treatment effect estimate will be biased in general for any of the aforementioned approaches.^1,3^ In practice, observed data cannot be used to determine whether data are MAR or MNAR, and, consequently, if estimates from CCA, MI or IPW analyses are likely to be biased.

Permutt & Li^4^ suggested a trimmed means (TM) estimator for RCTs where patients who drop out have more extreme (unobserved) outcome values than those who do not drop out (e.g., lower value dropout, when comparator group patients, not experiencing a benefit of treatment, leave the study early). Observations are ordered within each treatment group, with the missing observations assigned the lowest rank. Equal proportions of data are trimmed away from the lower end of both treatment group distributions and the TM treatment effect estimate is obtained from a linear regression of the exposure and outcome variables, using the remaining “trimmed” data.

The TM estimator will give an unbiased estimate of the true treatment effect, given two main assumptions - the location shift assumption and the strong MNAR assumption. The first specifies identical distributions of the treatment groups, with the only difference being a mean shift. The second restricts all dropout to the fraction that is trimmed away.^5^ Previous studies have performed a variety of simulations investigating the type 1 error and power of the TM estimator in a range of clinical scenarios^6^ and for various MNAR/MAR dropout patterns^5^. However, the biases that arise from violations of the strict TM assumptions and how to correct for these biases have yet to be established.

In this article, we derive formulae for the bias resulting from the violation of the location shift and strong MNAR assumptions for a normally distributed outcome, and illustrate, by means of simulations, the TM estimator bias for range of MNAR/MAR mechanisms. Additionally, we propose an adjustment to the estimator which relaxes the location shift assumption and leaves the estimator reliant only on the strong MNAR assumption and normality. For the purpose of this article, consistent with previous publications,^4,5,6^ we primarily consider worst value dropout. The principle, however, is equally applicable to higher value dropout, which may occur when patients leave the study perceiving themselves to be recovered, or where higher values indicate a worse response. We also primarily consider the case of 50% fixed trimming, but provide more general formulae for alternate trimming fractions. The TM approach is illustrated in an application to the CoBalT randomized controlled trial (registration ISRCTN38231611), which compares the effectiveness of <u>co</u>gnitive <u>b</u>ehaviour<u>al</u> therapy (CBT) as an adjunct to pharmacotherapy versus usual care in patients with treatment resistant depression. We have developed an R package ‘tmsens’ for performing a TM regression and conducting a TM estimator sensitivity analysis, available from https://github.com/dea-hazewinkel/tmsens.

## 2 Methods

### 2.1 Notation

Consider a clinical trial with *n* subjects, with a continuous outcome, *Y*, with patients randomized to receive an active treatment (*R* = 1) or comparator (*R* = 0). Let *y*_*ij*_ denote the observed outcome for patient *i* randomized to treatment group *j*, where *i* = 1, …, *n*_*j*_ and *j* = 0, 1. Let 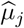 be the population mean for arm *j*, with *µ*_*j*_ = 𝔼 [*Y* | *R* = *j*], and its estimate, *µ*_*j*_, obtained from the corresponding sample mean:

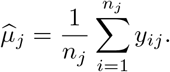

When there are no missing outcomes in the trial, each *µ*_*j*_ is an unbiased estimate of 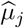 Let *β* denote the true treatment effect, given by the difference in treatment group means:

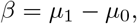

where *β* is estimated by 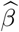:

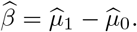

When the trial contains missing outcomes, we define a missing indicator *M*, which equals 0 if *y*_*ij*_ is observed and 1 if *y*_*ij*_ is not observed. Let 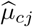 be the mean of all observed values for group *j*, given by 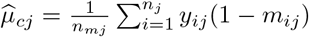, with 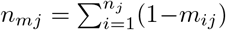 and *m*_*ij*_ being subject *i*’s missingness indicator. Then 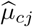 estimates the population complete case mean *µ*_*cj*_ = 𝔼 [*Y* |*R* = *j, M* = 0] and the complete case estimand of the treatment effect is given by

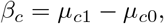

with *β*_*c*_ estimated by

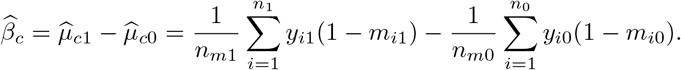

### 2.2 Trimmed means estimator

We first define the population trimmed mean, *µ*_*tj*_, in the absence of dropout (*m*_*ij*_ = 0 ∀ *i*). For treatment group *j, µ*_*tj*_ is given by the expected value of all observations exceeding the quantile *F* ^−1^(*p*):

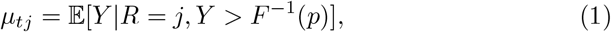

where *p* is the proportion of outcomes trimmed away from the lower end of the distribution of each group *j* (e.g., for *p* = 0.25, the bottom 25% of the distribution would be removed). Then *µ*_*tj*_ is estimated by

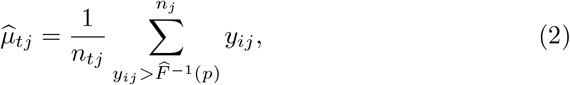

with *n*_*tj*_ being the sample size after trimming (*n*_*tj*_ = ⌈ *n*_*j*_ (1 − *p*) ⌉, with ⌈ ⌉the ceiling function).

Let *β*_*t*_ denote the TM effect estimand, given by the difference in population trimmed means (*β*_*t*_ = *µ*_*t*1_ − *µ*_*t*0_), estimated by 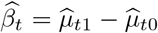. If the outcomes within each treatment group are normally distributed with underlying mean *µ*_*j*_ and variance 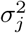, then *µ*_*tj*_ = 𝔼 [*Y* |*R* = *j, Y* ≥ *σ*_*j*_Φ^−1^(*p*) + *µ*_*j*_]. If the treatment group standard deviations (SDs) are equal (*σ*_1_ = *σ*_0_), *C* = *σ*_*j*_Φ^−1^(*p*) is common across treatment groups and the population TM effect, *β*_*t*_, is identical to the population mean difference *β*, since

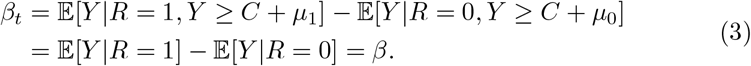

This property will also hold in absence of normality, if the outcome distributions for each group are identical in shape and differ only by a mean shift.

We now consider the TM estimator in the presence of dropout. As the estimator assumes worst value dropout, all missing values (*m*_*ij*_ = 1) are assigned a value smaller than the worst observed outcome:

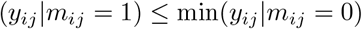

The precise value is unimportant as long as the above inequality holds. The trimmed mean is obtained by taking the average of all observations exceeding the quantile of the trimming proportion, *p*, as in (2), where *p* must now be equal to or exceed the largest observed dropout proportion across the groups:

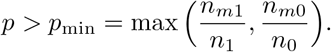

If there is no dropout in the trimmed fraction used to estimate *µ*_*t*_ (the “strong MNAR assumption”), 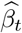 is an unbiased estimator for the TM effect *β*_*t*_. In addition, if the group variances are equal (the “location shift assumption”), *β*_*t*_ is unbiased for *β* (3). Previously, it has been stressed that the TM estimator estimates an unique estimand - the mean difference of the best X% patients in each treatment group, making it hard to compare with other approaches^5,6^. In order to identify this TM-specific estimand, only the strong MNAR assumption is required. Under the additional constraint of the location shift assumption, the TM effect becomes an unbiased estimator of the population mean difference as shown in (3).

We now examine the two sources of bias in the TM estimator. Let *B*_*t*_ be the total bias for the TM estimator, representing the deviation of the TM estimate, 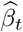, from the population mean difference, *β*:

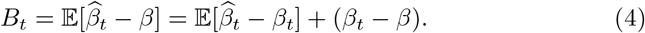

The first component results from the violation of the strong MNAR assumption, the second from violation of the location shift assumption. Supplementary Figure S1 (Appendix A) illustrates the TM and CCA estimator behaviours across four scenarios with varying dropout patterns and equal and unequal treatment arm SDs. Box 1 summarizes the terminology used throughout this paper.

### 2.3 The location shift assumption

We now derive expressions for the bias resulting from the violation of the location shift assumption, *B*_*tLS*_, for the case of 50% trimming, with results for a more general trimming fraction, *p*, derived in Appendix B. We assume that the strong MNAR assumption is satisfied and the outcomes are normally distributed for each treatment arm. Then, 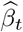 is an unbiased estimator for *β*_*t*_ and the total bias (4) is given by

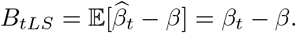

We define the 50% trimmed left-truncated mean (Appendix B):

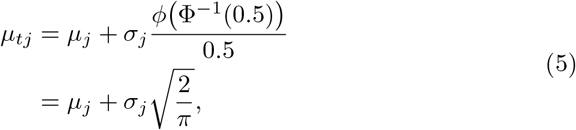

Then, the 50% trimmed population mean difference is given by

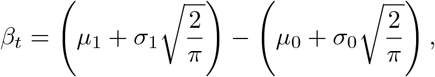

with the bias, *B*_*tLS*_, resulting from unequal SDs:

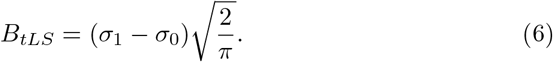

The TM estimator bias, *B*_*tLS*_ (6), can alternatively be expressed as a function of the trimmed fraction SDs, rather than of the unobserved full sample SDs (Appendix C):

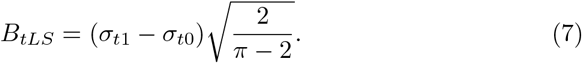

From both *B*_*tLS*_ bias formulae, (6) and (7), it is apparent that the estimator is unbiased given equal treatment group SDs, with the latter notation (7) making it explicit that equality of trimmed fraction SDs suffices. This property underlies the adjustment to the TM estimator outlined in Section 2.5, which relaxes the location shift assumption, allowing for different treatment group SDs.

#### BOX 1

**List of terms used throughout this paper**

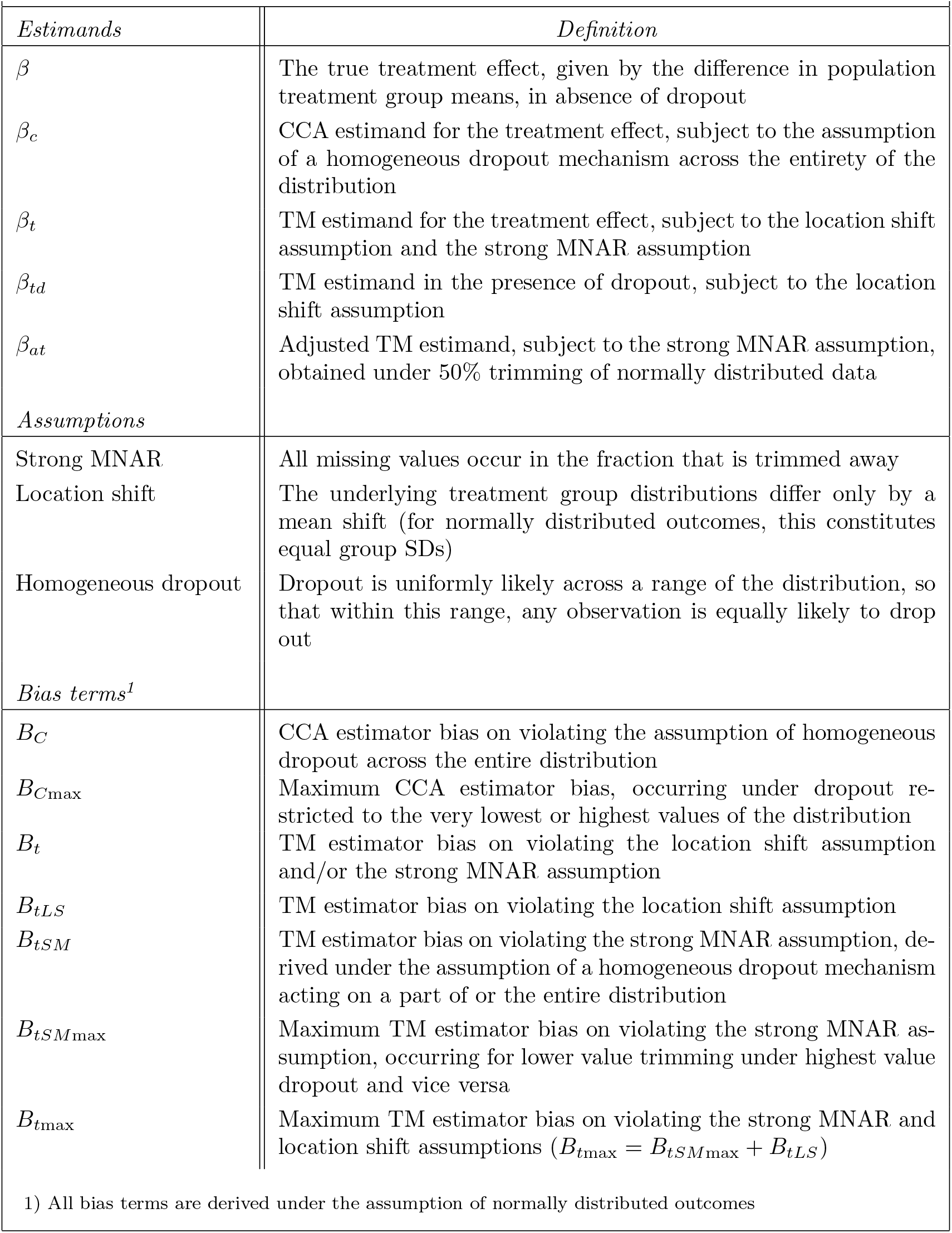

### 2.4 The strong MNAR assumption

We now derive expressions for the bias resulting from the violation of the strong MNAR assumption, *B*_*tSM*_, for dropout in the comparator group, with results for dropout in both groups derived in Appendix E. We examine violation due to a specific homogeneous dropout mechanism, where, in a given part of the distribution, any observation is equally likely to be selected for dropout.

We assume that the location shift assumption is satisfied and the outcomes are normally distributed in each treatment arm, prior to dropout. Then, *β*_*t*_ is an unbiased estimator for *β* and the total bias (4) reduces to

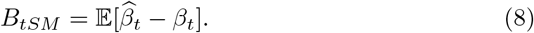

We define a new population parameter, *β*_*td*_, which gives the population TM effect for a scenario with a normally distributed treatment group and a comparator group no longer normally distributed due to dropout. Then, *β*_*t*_ unbiasedly estimates *β*_*td*_, the TM effect, and the bias, *B*_*tSM*_ (8), can be written as

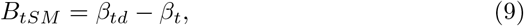

and, with *β*_*t*_ = *µ*_*t*1_ − *µ*_*t*0_ and *β*_*td*_ = *µ*_*t*1_ − *µ*_*td*0_, simplified to

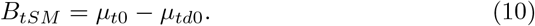

While *µ*_*td*0_ is the trimmed mean of a population with non-normally distributed outcomes, we can express it in terms of a normal distribution, under the assumption of a homogeneous dropout mechanism. Consider a normal distribution with lower bound, Φ^−1^(0) and upper bound, Φ^−1^(1), and let *f*_*u,v*_ denote the fraction of this distribution, so that *f*_0,1_ = 1. Let *f*_0,*c*_ denote the fraction affected by homogeneous dropout, with *c* ≤ 1, and Φ^−1^(*c*) its upper bound, so that each value in *f*_0,*c*_ is equally likely to drop out, with a given probability *a*:

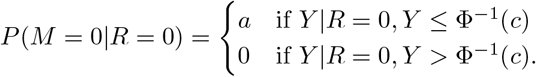

Further, let *f*_0,0.5_ denote the fraction of the distribution that is trimmed away under 50% trimming, *f*_0.5,1_ the trimmed fraction used for estimation, and *f*_0.5,*c*_ the fraction of the distribution for which the trimmed fraction, *f*_0.5,1_, is affected by dropout (in violation of the strong MNAR assumption), see Figure 1.

**Figure 1:**
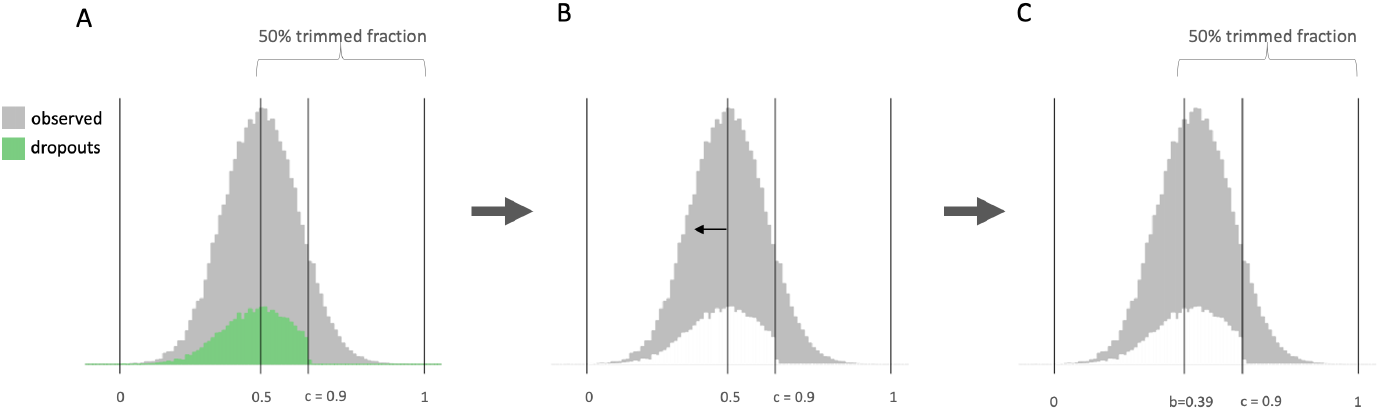
Schematic illustration of the dropout bias mechanism under assumption of homogeneity, for 50% trimming, and 20% dropout (*p*_*d*_ = 0.2) spread across 90% of the distribution (*f*_0,*c*_= 0.9). A) dropouts (green) and observed patients (grey) for normally distributed outcomes of a given group. The 50% trimmed fraction is given by *f*_0.5,1_, *f*_0.5,*c*_ gives the fraction affected by dropout, *f*_*c*,1_ the fraction unaffected by dropout, and *f*_0,0.5_ the fraction that is trimmed away. B) Under dropout, *f*_0.5,1_ lacks the observations to make up the 50% trimmed fraction, and the lower boundary, 0.5, shifts downwards in compensation. C) Under dropout, the 50% trimmed fraction is given by *f*_*b*,1_. The size of the shift is calculated with (11), giving, for this example, *b* = 0.39.

The 50% trimmed mean is estimated by taking the top half of the distribution under assumption of worse value dropout. When the strong MNAR assumption is satisfied, the trimmed mean is given by *µ*_*t*_ = *µ*_0.5,1_. When the assumption is violated, the fraction *f*_0.5,1_ no longer contains a sufficient number of observations, and a larger fraction of the distribution, *f*_*b*,1_, is taken, with *b* < 0.5 and the trimmed mean given by *µ*_*td*_ = *µ*_*b*,1_. The shift from 0.5 to *b* is a function of the dropout proportion, *p*_*d*_, the dropout spread, *f*_0,*c*_, the trimming proportion, *f*_0,0.5_ = *p*, and *f*_0.5,*c*_ = *f*_0,*c*_ − *p*, with

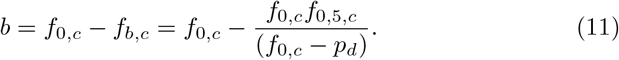

Using this notation, the bias of the TM estimator in (10) can be expressed as

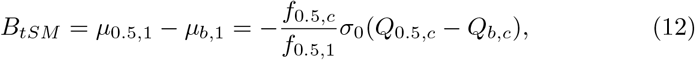

with, for example, *Q*_*bc*_ = (*f* Φ^−1^ ((*c*) − *f* Φ^−1^(*b*) /(*c* − (*b*)). The full derivation of (12) is given in Appendix D. *B*_*tSM*_ (12) is a function of the size of the trimmed fraction (*f*_0.5,1_), the fraction affected by dropout (*f*_0.5,*c*_), the SD of the affected group (*σ*_0_), and the magnitude of the shift from 0.5 to *b* (11), which is affected by the dropout proportion (*p*_*d*_). When the strong MNAR assumption is satisfied, no shift occurs, since *b* = 0.5, so that *Q*_0.5,*c*_ = *Q*_*b,c*_, and *B*_*tSM*_ reduces to 0.

Figure 1 illustrates the strong MNAR bias mechanism for the case of 50% trimming, with 20% dropout (*p*_*d*_ = 0.2) spread across 90% (*f*_0,*c*_ = 0.9) of the distribution. From (11), we obtain 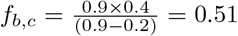 and *b* = 0.9 − 0.51 = 0.39.

With (12), we calculate *Q*_0.5,*c*_ = 0.59 and *Q*_*b,c*_ = 0.34. Specifying *σ*_0_ = 1, the bias, *B*_*tSM*_, is then given by 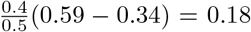. The bias is positive, and the treatment effect is overestimated, resulting from underestimation of the comparator group mean due to dropout in the trimmed fraction.

The bias (12) is defined under the assumption of a homogeneous dropout mechanism. In reality, dropout is unlikely to be homogeneous. In any scenario, however, where the TM approach can be considered an appropriate estimator, there should be reason to believe that the dropout comprises on average lower - worse - outcome values. Then, homogeneous dropout can be considered a worst-case scenario and the bias calculated under this assumption will serve as an upper bound for the bias. This property is illustrated in supplementary Figure S2 (Appendix F).

Unlike the TM estimator, the CCA estimator will be biased under the strong MNAR assumption. We define this bias, *B*_*C*_, for the case of dropout in the comparator group, with results for dropout in both groups derived in Appendix G. As for the TM estimator, we derive the bias under the assumption of a homogeneous dropout mechanism and a normally distributed outcome, with *B*_*C*_ given by

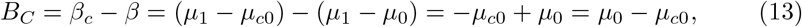

and

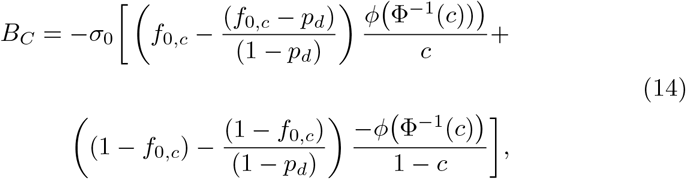

with *f*_0,*c*_ the dropout spread, *c* its upper bound, and *p*_*d*_ the dropout proportion. We fully derive (14) in Appendix G.

In the absence of dropout (*p*_*d*_ = 0), and for dropout spread homogeneously across the entire distribution (*c* = *f*_0,*c*_ = 1), *B*_*C*_ is 0. The CCA estimate will be maximally biased under dropout restricted to the very lowest or highest values of the distribution, the former resulting in overestimation of the complete case mean, *µ*_*cj*_, the latter in its underestimation. We can write the maximum bias for a given treatment group *j* under high or low value dropout as

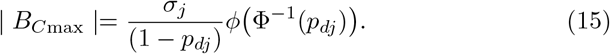

The full derivation of (15) is given in Appendix H.

The dropout mechanism, under which maximum CCA bias is achieved, can be defined in terms of a threshold selection model. Consider dropout of the *p*_*d*_ highest values, so that all observations exceeding the (1 − *p*_*d*_)’th quantile of the outcome distribution are unobserved. Let us define this threshold as *t*_*H*_ = *µ*_*j*_ + *σ*_*j*_Φ^−1^(1 − *p*_*dj*_) = *µ*_*j*_ + *σ*_*j*_Φ^−1^(*p*_*sj*_), so that

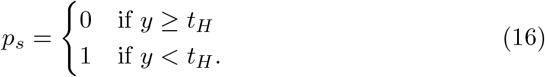

In the broader context of selection models in general, Copas and Jackson^7^ defined a bias limit, which is equivalent to the maximum CCA estimator bias that we derive here (15). We extend Copas and Jackson’s bias limit to the TM estimator, and define the maximum possible TM bias, which, for the case of lower value trimming, occurs under highest value dropout. For the comparator group, the maximum bias resulting from dropout is then given by

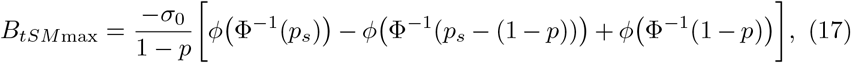

with the total maximum bias, *B*_*t*max_, accounting for the potential violation of the location shift assumption, given by

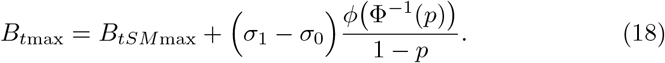

Full derivations of (17, 18) are given in Appendix I, alongside expressions for maximum bias under dropout in the treatment group. Appendix J describes a simple simulation illustrating the maximum CCA and TM estimator biases under dropout assumption violations.

### 2.5 Adjusted estimator

Here, we define an adjusted estimator which relaxes the location shift assumption. Under the assumption of normally distributed outcomes, the 50% trimmed fractions will have half-normal distributions. The SD of a given fraction can be adjusted by mirroring this half-normal distribution, and rescaling this now complete, if artificial, normal distribution, using the full sample SD of the other group. The latter SD will either be observed, in the absence of dropout, or can alternatively be extrapolated from the observed fraction SD, using the underlying properties of normality. The adjustment can be performed on either treatment groups, but we illustrate the procedure for the comparator group. Let *µ*_*t*0_ be the unadjusted 50% comparator trimmed mean, with

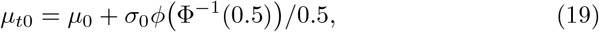

and *µ*_*at*0_ the adjusted comparator trimmed mean, with

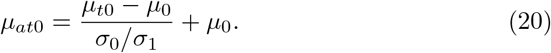

Substituting (19) for *µ*_*t*0_ in (20) then gives the comparator group trimmed mean under the treatment group SD, *σ*_1_. From the adjusted comparator trimmed mean, *µ*_*at*0_ (20), and the unadjusted treatment trimmed mean, *µ*_*t*1_ (5), we obtain the population adjusted TM estimate, under comparator group rescaling, *β*_*at*0_, as

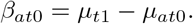

Equivalently, we can obtain the population adjusted TM estimate, under treatment group rescaling, *β*_*at*1_ = *µ*_*at*1_ − *µ*_*t*0_, with *µ*_*at*1_ defined analogously to (20). As for the unadjusted estimator, violation of the strong MNAR assumption will bias the estimate, with violation in the comparator group resulting in an overestimation of the treatment effect, and the converse true for the treatment group. We show in simulation (Section 3, Table 1) that this assumption is stronger for the adjusted estimators than for the unadjusted estimator, with the greatest bias observed when rescaling the group for which the strong MNAR assumption is most violated, and recommend that the adjustment be applied to the group for which the assumption is most plausible. In Appendix K, expressions are derived for the adjusted estimator bias resulting from strong MNAR assumption violation in either or both treatment groups, under comparator group rescaling and under treatment group rescaling.

**Table 1:** Estimated treatment effects 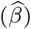 and corresponding biases 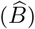 for the CCA 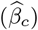, TM 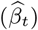 and adjusted TM 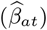 estimators, with the latter obtained when performing the adjustment on the comparator group 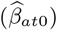 and the treatment group 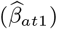. Mean and standard deviation (SD) estimates for *S* = 1000 simulations are reported. The TM estimands are estimated under 50% trimming of normally distributed outcomes (*N* = 1000), for equal (a) and unequal (b) treatment group SDs, true treatment effect *β* = 0.5, and 20% comparator group dropout. Four scenarios are considered: dropouts restricted to 1) the lowest 20% of the distribution (*f*_*ac*_ = 0.2), 2) the lowest 50%, 3) the lowest 75% and 4) left unrestricted.

| Dropout spread | | $\hat{\beta}_c$ | $\hat{\beta}_t$ | $\hat{\beta}_{at1}$ | $\hat{\beta}_{at0}$ | $\hat{B}_{tLS}$ | $\hat{B}_{tSM}$ | $\hat{B}_t$ | $\hat{B}_{at1}$ | $\hat{B}_{at0}$ |
| --- | --- | --- | --- | --- | --- | --- | --- | --- | --- | --- |
| 1) $f_{0,c} = 0.2$ | $\hat{\beta}/\hat{B}$ | 0.15 | 0.50 | 0.50 | 0.50 | 0.00 | 0.00 | 0.00 | 0.00 | 0.00 |
| | $\overline{SD}$ | 0.02 | 0.02 | 0.03 | 0.03 | 0.02 | 0.02 | 0.02 | 0.03 | 0.03 |
| 2) $f_{0,c} = 0.5$ | $\hat{\beta}/\hat{B}$ | 0.30 | 0.50 | 0.50 | 0.50 | 0.00 | 0.00 | 0.00 | 0.00 | 0.00 |
| | $\overline{SD}$ | 0.02 | 0.02 | 0.03 | 0.03 | 0.02 | 0.02 | 0.02 | 0.03 | 0.03 |
| 3) $f_{0,c} = 0.75$ | $\hat{\beta}/\hat{B}$ | 0.40 | 0.56 | 0.64 | 0.70 | 0.00 | 0.06 | 0.06 | 0.14 | 0.20 |
| | $\overline{SD}$ | 0.02 | 0.02 | 0.03 | 0.03 | 0.02 | 0.02 | 0.02 | 0.03 | 0.03 |
| 4) $f_{0,c} = 0.1$ | $\hat{\beta}/\hat{B}$ | 0.50 | 0.69 | 0.77 | 0.81 | 0.00 | 0.19 | 0.19 | 0.27 | 0.31 |
| | $\overline{SD}$ | 0.02 | 0.02 | 0.03 | 0.03 | 0.02 | 0.02 | 0.02 | 0.03 | 0.03 |

(b) $\sigma_0 = 1.5, \sigma_1 = 1$
| Dropout spread | | $\hat{\beta}_c$ | $\hat{\beta}_t$ | $\hat{\beta}_{at1}$ | $\hat{\beta}_{at0}$ | $\hat{B}_{tLS}$ | $\hat{B}_{tSM}$ | $\hat{B}_t$ | $\hat{B}_{at1}$ | $\hat{B}_{at0}$ |
| --- | --- | --- | --- | --- | --- | --- | --- | --- | --- | --- |
| 1) $f_{0,c} = 0.2$ | $\hat{\beta}/\hat{B}$ | -0.03 | 0.10 | 0.50 | 0.50 | -0.40 | 0.00 | -0.40 | 0.00 | 0.00 |
| | $\overline{SD}$ | 0.03 | 0.03 | 0.03 | 0.03 | 0.03 | 0.03 | 0.03 | 0.03 | 0.03 |
| 2) $f_{0,c} = 0.5$ | $\hat{\beta}/\hat{B}$ | 0.20 | 0.10 | 0.50 | 0.50 | -0.40 | 0.00 | -0.40 | 0.00 | 0.00 |
| | $\overline{SD}$ | 0.03 | 0.03 | 0.03 | 0.03 | 0.03 | 0.03 | 0.03 | 0.03 | 0.03 |
| 3) $f_{0,c} = 0.75$ | $\hat{\beta}/\hat{B}$ | 0.34 | 0.19 | 0.70 | 0.82 | -0.40 | 0.09 | -0.31 | 0.20 | 0.32 |
| | $\overline{SD}$ | 0.03 | 0.03 | 0.04 | 0.04 | 0.03 | 0.03 | 0.03 | 0.04 | 0.04 |
| 4) $f_{0,c} = 1$ | $\hat{\beta}/\hat{B}$ | 0.50 | 0.39 | 0.91 | 0.96 | -0.40 | 0.29 | -0.11 | 0.41 | 0.46 |
| | $\overline{SD}$ | 0.03 | 0.03 | 0.03 | 0.03 | 0.03 | 0.03 | 0.03 | 0.03 | 0.03 |

## 3 A simulation study

Consider a clinical trial on *n* = 1000 subjects randomized to either a treatment (*j* = 1) or comparator (*j* = 0) arm, with *n*_1_ = *n*_0_ = 500. We assume the outcomes are normally distributed, with a true treatment effect *β* = 0.5, and 20% dropout in the comparator group. Tables 1a and 1b give the mean treatment effect estimates and bias across *S* = 1000 simulations for the CCA estimator and the TM estimator under 50% trimming, for four scenarios. In the first three, dropout is restricted to the lowest 20%, 50% and 75%, respectively, of the comparator arm outcome distribution, and left completely unrestrained in the fourth scenario.

In the first two scenarios, the strong MNAR assumption is satisfied and we observe in Table 1a, for equal treatment SDs (*σ*_1_ = *σ*_0_ = 1), that the TM estimate, 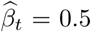, is unbiased, while the CCA estimate, 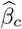, is biased towards the null. On violating the strong MNAR assumption, this bias decreases, with 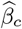 being unbiased under random dropout (*f*_*ac*_ = 1). We observe the same pattern for *σ*_0_ = 1.5 and *σ*_1_ = 1 (Table 1b), with the CCA bias now larger due to the higher *σ*_0_, which acts multiplicatively on the bias (14). Under unequal SDs, the location shift assumption is violated, and the TM estimates are biased towards the null. For the first two scenarios, the total bias, *B*_*t*_, is equal to *B*_*tLS*_, while in scenarios 3 and 4, *B*_*t*_ is given by the sum of *B*_*tLS*_ and *B*_*tSM*_, with the latter biasing the TM estimates away from the null. As for the CCA estimator, the increased comparator group SD results in a larger *B*_*tSM*_ bias in Table 1b.

The adjusted TM estimates are given by 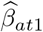 and 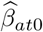, for rescaling applied to the treatment and comparator group, respectively. In the first two scenarios, the strong MNAR assumption is satisfied, and both estimators estimate an unbiased treatment effect, *β* = 0.5, under unequal SDs (Table 1b). For scenarios 3 and 4, however, both are biased away from the null, with bias components, 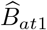 and 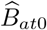, bigger than the unadjusted estimator bias, 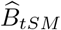. The greatest bias is observed for 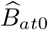, a consequence of performing the adjustment on the group for which the strong MNAR assumption is violated.

Examples of bias calculations for the CCA and (un)adjusted TM estimators are given in Appendix L. Appendix M describes an additional simulation, for trimming fractions, *p*, other than 50%, illustrating the bias trade-off between the *B*_*tLS*_ and *B*_*tSM*_ bias components. Appendix N provides R code for obtaining adjusted and unadjusted TM estimates for a simulated dataset.

## 4 An application to the CoBalT randomized clinical trial

The CoBalT trial^9,10^ was a multicentre trial investigating the effect of CBT as an adjunct to pharmacotherapy versus usual care (UC) in 469 patients aged 18-75 with treatment-resistant depression. The primary outcome was the BDI-II score - a self-completed measure of depressive symptoms, with higher values indicating greater depression. Long-term treatment effect was assessed by a repeated measures intention-to-treat analysis (*N* = 396), using outcomes at 6 months, 12 months and 3-5 years (average 46 months) and adjusted for BDI-II at baseline, treatment centre and minimization variables (previously prescribed antidepressants, presence of a counselor at the general practice, and duration of current depressive episode at baseline). The repeated measures analysis showed a beneficial effect of CBT versus UC of −4.7 (95% CI: −6.4,-3.0) with a mean difference of −3.6 (95% CI: −6.6,-0.6) at 46 months for 136 and 112 patients in the CBT and UC group, respectively.^10^

### 4.1 Missing data

Just over half of the initially recruited patients were observed at the final follow-up, with greater dropout in the UC group than the CBT group (Table 2). Dropout in the UC group appears to be unrelated to the most recent depression score, whereas in the CBT group, dropout was more common in those with higher recent depression scores (Table 3). The CBT group SDs are higher, most noticeably for patients missing at 46 months, suggesting that the unobserved full sample CBT SD at 46 months may be higher than the UC group SD. In Supplementary Figure S3 (Appendix O), histograms of the BDI-II score distributions are given, across time, for the UC group, the CBT group and their combined total. We observe that the distributions are moderately right-skewed and non-normal.

**Table 2:** Counts (*N*) and percentages (%) of patients with an observed BDI-II score at baseline, 6 months, 12 months, and 46 months, for the usual care group (UC), the CBT treatment group (CBT), and both groups

|  | UC |  | CBT |  | Total |  |
| --- | --- | --- | --- | --- | --- | --- |
|  | N | % | N | % | N | % |
| Base | 235 | 100 | 234 | 100 | 469 | 100 |
| 6 mo. | 213 | 90.6 | 206 | 88.0 | 419 | 89.3 |
| 12 mo. | 198 | 84.3 | 197 | 84.2 | 395 | 84.2 |
| 46 mo. | 112 | 47.7 | 136 | 58.1 | 248 | 52.9 |

**Table 3:** Mean BDI-II measurements and SDs at baseline, 6 months, 12 months and 46 months for the usual care group (UC), the CBT treatment group. Patients missing and observed at 46 months are compared for all time points, with mean differences and 95% confidence intervals provided.

|  | Usual care (UC) |  |  | CBT |  |  |
| --- | --- | --- | --- | --- | --- | --- |
|  | Observed<sup>a</sup> | Missing<sup>a</sup> | Difference<sup>b</sup> | Observed<sup>a</sup> | Missing<sup>a</sup> | Difference<sup>b</sup> |
| Base | 30.9 (10.3) | 32.7 (11.3) | -1.8 (-4.6,1.0) | 30.4 (9.3) | 33.6 (11.7) | -3.2 (-5.9,-0.5) |
| 6 mo. | 24.6 (13.1) | 24.4 (13.2) | 0.2 (-3.3,3.8) | 17.4 (13.6) | 21.8 (14.9) | -4.5 (-8.5,-0.4) |
| 12 mo. | 21.8 (13.3) | 21.5 (12.4) | 0.2 (-3.4,3.9) | 15.8 (12.9) | 19.5 (15.8) | -3.8 (-7.9,0.4) |
| 46 mo. | 23.4 (13.2) |  |  | 19.2 (13.8) |  |  |

a) Given are mean and standard deviation (SD)
b) Given is the mean difference (observed - missing) with 95% confidence interval

### 4.2 Analysis details

Using the maximum CCA bias formula (15) from Section 2.4, we would expect the treatment effect to either under- or over-estimate the treatment effect by approximately 10 units of the BDI-II score. This is a wide bound, obtained without taking into account information available from the data and motivating the use of a more precise sensitivity analysis. To this end, we employed the TM approach and two MI models as a sensitivity analysis for the treatment effect at 46 months, which is estimated using linear regression adjusting for treatment centre, BDI-II at baseline and various minimization variables (previously described antidepressants, presence of a counselor at the general practice, duration of current depressive episode at baseline). Adaptive trimming was employed due to the high dropout proportion observed, with 52.3% of the highest values trimmed away in both groups. Confidence intervals were obtained using a permutation-based approach.^4^ We considered only the unadjusted TM estimator, as the adjusted estimator (Section 2.5), which adjusts for unequal treatment group SDs, is strongly reliant on normality. The imputation models included auxiliary variables: baseline variables associated with missing BDI-II at any time of follow-up, and various measures of depression and anxiety (BDI-II, PHQ-7, GAD-7, SF-12). Two MI models were considered, with the first using only depression/anxiety information available at baseline (MI-baseline), the second including intermediate outcome measurements at all follow-up times.

To interpret the sensitivity analysis results, we used the bias formulae derived in Sections 2.3 and 2.4 to establish CCA and TM estimator bias directions under different dropout scenarios. Additionally, we calculated the expected TM estimator bias of the CBT treatment effect in the CoBalT data, for three variations of the most plausible dropout scenario. The location shift assumption bias and strong MNAR bias are both functions of the full sample SDs, which remain unobserved. Subject to the specified dropout mechanism, the observed dropout proportions, and under assumption of normally distributed outcomes, these can be inferred from the observed SDs, and used to calculate the relevant bias terms (derivations and R code in Appendix P).

### 4.3 Plausibility of assumptions for CCA, MI and TM estimators

The validity of each method rests on the underlying estimator assumptions about data characteristics, assumptions which cannot (usually) be tested. We used the bias formulae derived in Sections 2.3 and 2.4 to compare TM and CCA estimator behaviour across four plausible dropout scenarios Figure 2).

**Figure 2:**
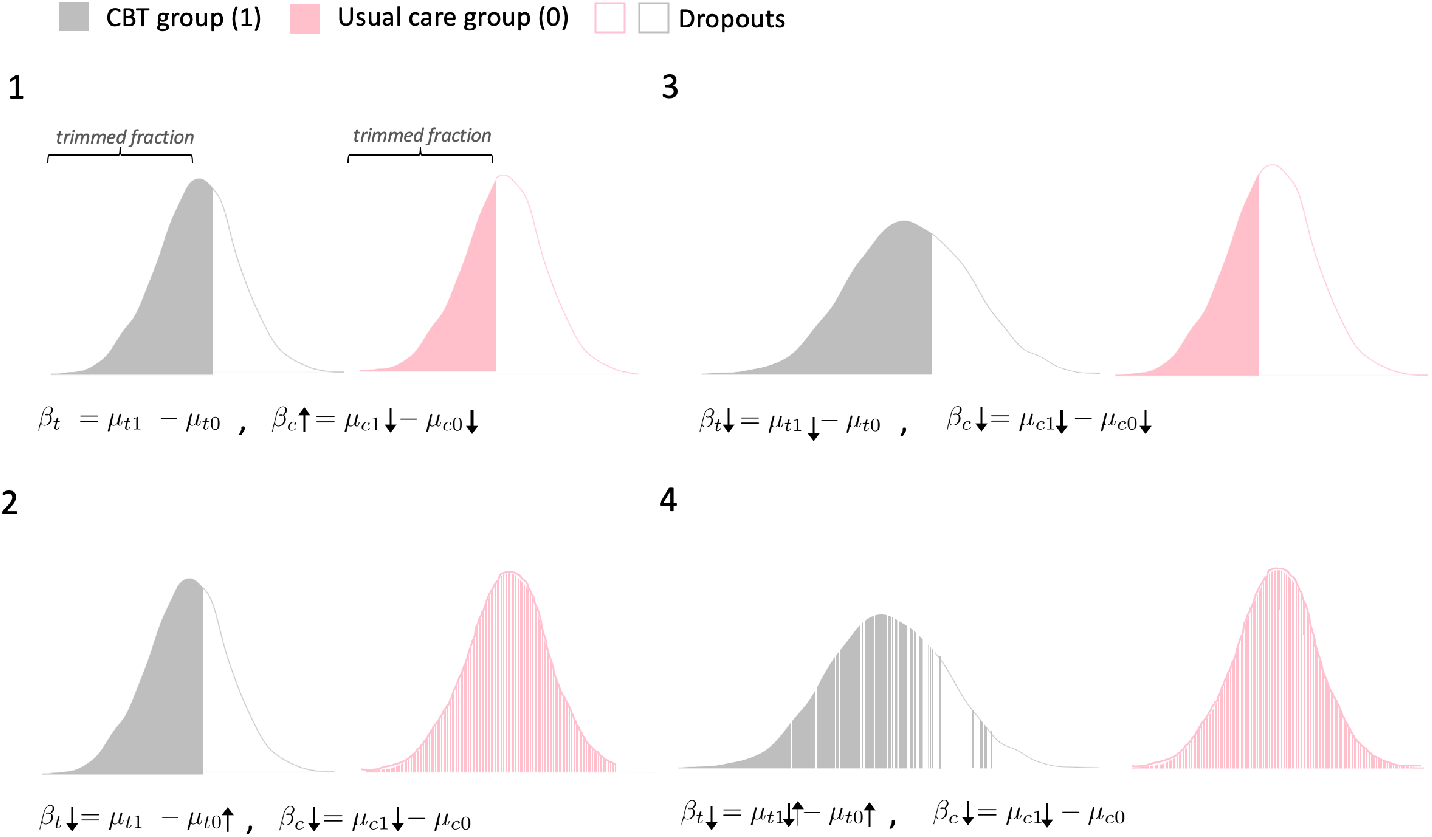
Trimmed means (TM) and complete case analysis (CCA) estimators across four scenarios with varying dropout patterns and treatment arm distributions, for 52.3% dropout in the usual care group (pink), 41.9% dropout in the CBT group (grey) and higher value 52.3% trimming. 1) The location shift assumption and strong MNAR assumption are satisfied, with equal treatment group SDs and strictly higher value dropout. 2) The location shift assumption is satisfied, but the strong MNAR assumption is violated in the usual care group. 3) The location shift assumption is violated, with *σ*_1_ > *σ*_0_, and the strong MNAR assumption is satisfied. 4) The location shift assumption is violated with *σ*_1_ > *σ*_0_, and the strong MNAR assumption is violated in the usual care group and to a lesser degree in the CBT group.

The CCA estimator will be biased if data are MNAR. Dropout of more depressed patients in the CBT group will result in underestimation of the CBT complete case mean, with the CCA estimator consequently overestimating the beneficial effect of CBT (Figure 2.2).

The TM estimator will be unbiased if the location shift assumption and strong MNAR assumption hold (Figure 2.1). From the CoBalT data, we suspect dropout of patients with more severe depressive symptoms in the CBT group (higher value dropout) and (approximately) homogeneous (MCAR) dropout in the UC group. This will, under higher value trimming, lead to the inclusion of too many high values in the trimmed fraction, and consequently to overestimation of the UC trimmed mean and the treatment effect (Figure 2.2). Intermediate BDI-II measurements at earlier times suggest a comparatively higher SD for the CBT group, implying that the location shift assumption may be violated (Figure 2.3), with the larger spread of values leading to underestimation of the CBT trimmed mean and overestimation of the beneficial effect of CBT. Violation of the location shift assumption, with SD_CBT_ > SD_UC_, combined with violation of the strong MNAR assumption in the UC group, will exacerbate the bias, leading to a substantial overestimation of the CBT treatment effect. This is illustrated in Figure 2.4, where the bias is somewhat mitigated by allowing for the possibility that the strong MNAR assumption may also be violated, if to a much lesser extent, in the CBT group.

MI estimates will be unbiased if the data are MAR and the imputation model is correctly specified. Under higher-value dropout in the CBT group, the MI models will impute BDI-II values that are too low in the CBT group, but approximately correct in the UC group, resulting in underestimation of the CBT imputed mean. As with the CCA estimator, this will result in a bias away from the null, with the MI estimators overestimating the benefit of CBT treatment. The inclusion of auxiliary information (baseline and intermediate depression measures) in the MI models could potentially overcome part of the MNAR bias, resulting in estimates that are closer to the truth than CCA. In summary, our bias formulae suggest that under plausible assumptions, TM, MI and CCA will overestimate the treatment effect.

### 4.4 Results

The CCA treatment effect estimate for CBT of −3.89 (95% CI: −6.95,-0.83) is comparable to the one of the main CoBalT follow-up analysis^10^ (Table 4). Slightly smaller effects are estimated by the MI models, comparable to the attenuation observed in the MI sensitivity analysis of the CoBalT follow-up analysis.^10^ The TM estimator, in contrast, estimated a considerably bigger beneficial CBT treatment effect of −8.26 (95% CI: −11.20,-5.32). These results are in line with our expectations regarding the BDI-II score and dropout characteristics, detailed in Section 4.4.

**Table 4:** Treatment effect estimate 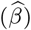) standard error (SE), and 95% confidence interval (95% CI), for the trimmed means (TM) estimator, complete case analysis (CCA), multiple imputation model with baseline outcome measurements (MI baseline) and multiple imputation model with intermediate outcome measurements (MI intermediate).

| | $\widehat{\beta}$ | SE | 95% CI |
| --- | --- | --- | --- |
| TM | -8.26 | 1.47 | (-11.2, -5.03) |
| CCA | -3.89 | 1.53 | (-6.92, -0.87) |
| MI (baseline) | -3.33 | 1.39 | (-6.07, -0.58) |
| MI (intermediate) | -2.56 | 1.37 | (-5.65, -0.08) |

From the CoBalT data, we have reason to suspect higher value dropout in the CBT group and approximately homogeneous dropout in the UC group. Table 5 takes a closer look at the TM estimator bias components calculated under three different assumptions of the dropout mechanism. In scenario A, we assume completely directional, highest value dropout in the CBT group, with all dropout values (41.9%) restricted to the top of the distribution, and entirely homogeneous dropout in the UC group, with the dropout (53.3%) spread equally across the distribution. In scenarios B and C, we make less stringent assumptions, with in scenario B dropout in the top 60% for the CBT group, and in scenario C dropout in the top 60% of the CBT group and top 80% of the UC group. We obtain a total bias, 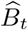 of −17.53, −11.09, and −7.1 for scenarios A, B and C, respectively, and calculate a bias-adjusted estimate, which can be interpreted as an upper bound for the treatment effect estimate, 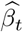, with the lower bound given by the TM estimate obtained from the data 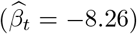. For all three scenarios, the CCA and MI estimates (Table 4) fall within the 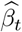 bounds, with the bias-adjusted estimate of scenario C most comparable, at −1.16. Our sensitivity analysis, comparing CCA, TM and MI, is consistent with a beneficial CBT treatment effect, but suggests that it may be more modest than indicated by the CCA estimator.

**Table 5:**
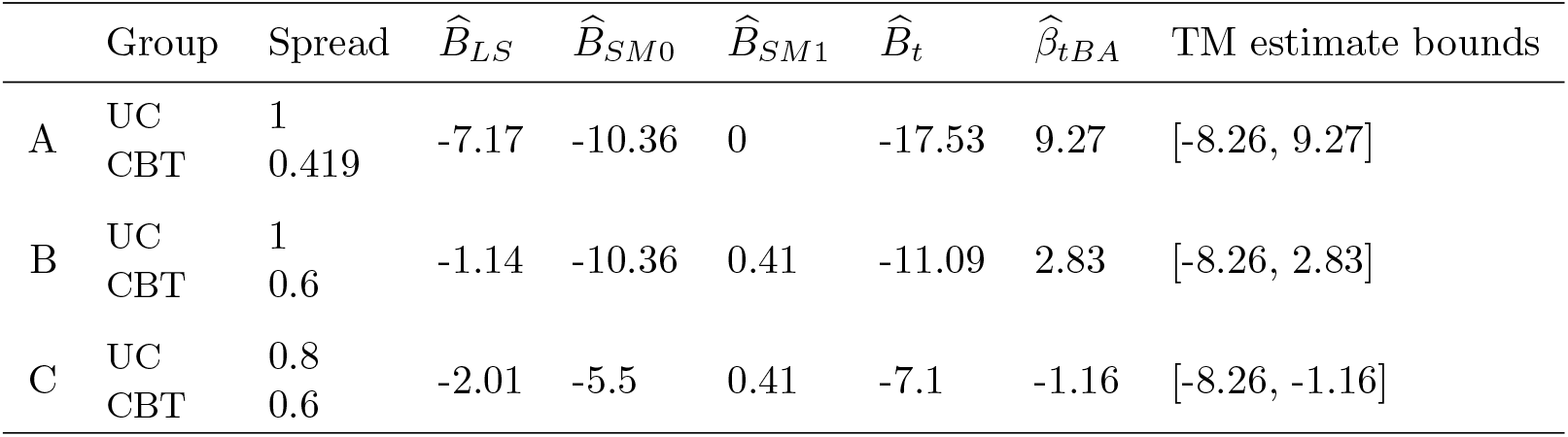
Trimmed mean estimator treatment effect bounds and bias components under different dropout assumptions. A) CBT group dropout restricted to the top 41.9% of the distribution and homogeneous UC group dropout; B) CBT group dropout in the top 60% of the distribution and homogenous UC group dropout; C) CBT group dropout in the top 60% of the distribution and UC group dropout in the top 80%. Shown are the bias components for violation of the location shift assumption 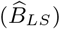, the strong MNAR assumption in the UC group 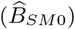, the strong MNAR assumption in the CBT group 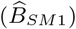, the total bias 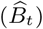, the bias adjusted estimate 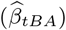, and the maximum bounds of the TM estimate, 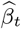.

| | Group | Spread | $\hat{B}_{LS}$ | $\hat{B}_{SM0}$ | $\hat{B}_{SM1}$ | $\hat{B}_t$ | $\hat{\beta}_{tBA}$ | TM estimate bounds |
| --- | --- | --- | --- | --- | --- | --- | --- | --- |
| A | UC | 1 | -7.17 | -10.36 | 0 | -17.53 | 9.27 | [-8.26, 9.27] |
|  | CBT | 0.419 |  |  |  |  |  |  |
| B | UC | 1 | -1.14 | -10.36 | 0.41 | -11.09 | 2.83 | [-8.26, 2.83] |
|  | CBT | 0.6 |  |  |  |  |  |  |
| C | UC | 0.8 | -2.01 | -5.5 | 0.41 | -7.1 | -1.16 | [-8.26, -1.16] |
|  | CBT | 0.6 |  |  |  |  |  |  |

## 5 Discussion

Our work extends existing TM literature by deriving formulae that can be used to calculate the resulting bias, establish bias direction, and also, under certain additional assumptions, calculate a limit for the maximum expected bias. In Section 4.3, we show how these formulae can be used to aid interpretation of sensitivity analyses. In addition to this, we describe an adjustment to the estimator that relaxes the location shift assumption.

Missing data is a common feature in RCTs and may result in biased inference, with any analysis relying on unverifiable assumptions about the relationship between observed and missing data. To increase confidence in the primary results, their robustness should be assessed by performing sensitivity analyses under a range of plausible alternative assumptions.^1,2,11,12^ While conducting such analyses in the presence of missing data is recommend practice, only a minority of affected trials report performing any kind of sensitivity analysis. Six review articles describing the handling of missing data in RCTs found that only 22% of the 649 trials reviewed reported some kind of sensitivity analysis.^13–18^ The majority retained the missingness assumptions of the original analysis, with only a small subset relaxing the original assumptions, and almost none considering a MNAR mechanism.^13^

Applying sensitivity analyses can be limited by the complexity of the methods, and a lack of transparency about the assumptions underlying each method. Two well-known approaches for dealing with incomplete data, which can be adapted to sensitivity analyses, are selection models (SM) and pattern mixture models (PMM). Selection models specify the relationship between outcome and the probability of being missing, while PMM define the outcome distribution for dropout across a range of missingness patterns.^2^ An example is the delta-adjusted PMM, which assumes that patients who drop out have a mean outcome that differs by a fixed amount, *delta*, to the ones who remain in study.^12^ This parameter, *delta*, is user-specified and typically informed by expert knowledge, in an attempt to characterize the unknown relationship between observed and unobserved data. However, choosing plausible values and distributions for sensitivity analysis parameters such as *delta* is far from straightforward.^19,20^ A simpler alternative is to specify some extreme scenarios, and, under these, re-estimate the main analysis results. An example of such an extreme scenario analysis is the combination approach of best-worst and worst-best case sensitivity analyses, in which beneficial outcomes are assigned to dropouts in one group and harmful ones to dropouts in the other group, and vice versa, yielding two estimates under opposing assumptions^21,22^. The TM estimator, operating under the assumption of strict directional dropout and requiring only the specification of the size of trimming fraction, can be considered part of this family of easily implemented extreme-case scenario estimators. Such simple bias-sensitivity analyses will often be sufficient to assess the robustness of inferences to potential bias sources^11^, but in the event of ambiguity, can be followed up with a more nuanced and fine-tuned sensitivity analysis (e.g. from the family of SM or PMM models).

While the TM estimator is simpler to implement than SM and PMM models, requiring no explicit specification of a missingness model or numerical sensitivity parameters, it is theoretically similar. The TM estimator can be thought of as a threshold SM, with the threshold value informed by the size of the trimming fraction, and with the probability of missing set to zero in the trimmed fraction and left unspecified in the part of the distribution that is trimmed away. A similar parallel can be drawn between TM and the delta-adjusted PMM, with the *delta* mean difference between observed and missing values implicit in the former and a direct result of the choice of trimming fraction. Under the strong MNAR assumption, this difference should be equal to or bigger than the difference between the mean of the trimmed fraction and the mean of the fraction that is trimmed away.

The TM estimator will give an unbiased estimate of the true treatment effect under the strong MNAR assumption and the location shift assumption. The TM estimator is only unbiased for the particular case of MNAR data where the trimmed fraction remains unaffected by dropout, and will be biased, even under the null, when outcomes are MAR. In practice, missing data will frequently be characterized by more than a single missingness mechanism (e.g., missing outcomes may be a mix of MAR and MNAR). Ocampo et al.^5^ sought to resolve this by combining the TM estimator with multiple imputation. This approach, however, relies on the assumption that missing values determined by MAR and MNAR can be distinguished using auxiliary information and its success will be heavily affected by the reliability and interpretation of the available information.

As assumptions underlying sensitivity analyses cannot be verified, it is important to know how different underlying data structures affect a given estimator. While previous studies have described the TM assumptions^4–6^, the biases arising from the violation of these strict assumptions have not previously been characterized. By quantifying the bias resulting from violation of these assumptions and establishing bias direction under a range of plausible data scenarios, we show how conclusions can be drawn from a sensitivity analysis, moving beyond a comparison of estimates. For example, in the CoBalT analysis (Section 4.4), the TM estimator, under very strict assumptions, estimated a beneficial effect of CBT treatment of approximately −8. However, when considering more plausible data generating scenarios and relaxing these assumptions, we obtained a bias-corrected estimate, which was much smaller.

In order to identify relevant sensitivity analyses, a framework, such as the one employed for the CoBalT analysis in Section 4.3, should be followed. Here, our initial bias results suggest that all estimators will be biased away from the null and overestimate the CBT treatment effect. The logical next step would involve identifying sensitivity analyses that might be biased in the opposite direction for a plausible data scenario. The TM estimator will be particularly useful in cases where it can be expected to be biased in the opposite direction to the CCA analysis. For example, if we had seen evidence of higher value dropout in both groups (i.e., previous depression being higher for those with missing outcome data in both UC and CBT groups) and evidence of similar SDs in the two arms, then we would expect the CCA estimator to underestimate the treatment effect given greater dropout in the comparator group, and overestimate the effect given higher dropout in the treatment group, while the TM estimator would remain unbiased for both.

Commonly applied sensitivity analyses employ complex models, for which bias quantification methods are not readily available. In contrast, the TM estimator is a simple approach, for which, as we show, analytic expressions for the bias due to missingness are available. In this paper, we show how the TM estimator can be used as a sensitivity analysis, by using bias formulae to interpret results in context of information available from the data, as we show in our CoBalT analysis. We recommend performing a TM sensitivity analysis when directional dropout is plausible, using the bias formulae to establish bias direction and interpret estimates.

## Data Availability

Data from the CoBalT trial is available on request from the authors

## Declarations

### Data Statement

Data from the CoBalT trial is available on request from the authors

### Funding statement

AH, TP and KT were supported by the Integrative Epidemiology Unit, which receives funding from the UK Medical Research Council and the University of Bristol (MC UU 00011/3). At the beginning of this project. JB’s research at the University of Exeter is funded by a UKRI Expanding Excellence in England (E3) award. KHW was supported by the Elizabeth Blackwell Institute for Health Research, University of Bristol and the Wellcome Trust Institutional Strategic Support Fund [204813/Z/16/Z] and is now affiliated to the Integrative Cancer Epidemiology Programme (ICEP), which is supported by a Cancer Research UK programme grant (C18281/A19169) and works within the Medical Research Council Integrative Epidemiology Unit (MC UU 00011/1-7). NW: The CoBalT trial was funded by the National Institute for Health Research Health Technology Assessment (NIHR HTA) programme (project number 06/404/02).

### Declaration of interest statement

The authors declare no conflicts of interest.

## Acknowledgments

The CoBalT trial was supported by the NIHR Biomedical Research Centre at University Hospitals Bristol and Weston NHS Foundation Trust and the University of Bristol. The views expressed are those of the author(s) and not necessarily those of the NIHR or the Department of Health and Social Care.

## Notes

### Competing Interest Statement

The authors have declared no competing interest.

### Clinical Trial

ISRCTN38231611

### Author Declarations

This paper made use of data from the CoBalT follow-up study. Ethical approval for the follow-up study was given by the National Research Ethics Service Committee West Midlands, Edgbaston (reference number 13/WM/0149). Research governance approvals were obtained from the relevant local Research Ethics Committees and Clinical Commissioning Groups or Health Boards covering the three study sites (Bristol, Exeter, and Glasgow). The protocol is available online.

