## Supplemental Material for "Sensitivity to missing not at random dropout in clinical trials: use and interpretation of the Trimmed Means Estimator"

### Appendix

A A visual illustration of location shift assumption bias and strong MNAR bias 3

B Location shift assumption bias for a given trimming fraction, $p$, as a function of full sample SDs 4

C Location shift assumption bias, as a function of trimmed fraction SDs, for a given trimming fraction, $p$, and for 50% trimming ($p = 0.5$) 5

D Strong MNAR assumption bias for dropout in the placebo group, for 50% trimming and for a given trimming fraction $p$ 7

E Strong MNAR assumption bias for dropout in both treatment groups 8

F A limited upper bound for the strong MNAR assumption bias 10

G CCA bias under homogeneous dropout in one or both treatment groups 11

H CCA bias for dropout restricted to the lowest part of the distribution 13

I Maximum bias of the TM estimator in relation to Copas' and Jackson's bias limit 14

J Simulation illustration: CCA and TM estimator bias for highest value placebo group dropout 16

K Adjusted estimator bias under strong MNAR violation in either or both treatment groups 18

M Trade-off of location shift bias and strong MNAR bias under different trimming proportions 27

N R code for obtaining unadjusted and unadjusted TM estimates 28

O Application using the CoBalT RCT: BDI-II score distributions across time and treatment groups 34

P Inferring the full sample SD from the SD observed under dropout, and calculating bias for the CoBalT application (R code) 34

### A A visual illustration of location shift assumption bias and strong MNAR bias

Supplementary Figure S1 compares the trimmed mean (TM) and complete case analyis (CCA) estimators across single realizations of four scenarios with varying dropout patterns and equal and unequal treatment arm standard deviations (SDs). In Figures S1A and S1B, the TM estimator is an unbiased estimator of the true treatment effect, $\beta$, under 50% trimming. The outcome values are normally distributed with underlying population means, $\mu_0$ and $\mu_1$, for placebo and treatment group, respectively, and equal SDs ($\sigma = \sigma_1 = \sigma_0$). Let $\widehat{\mu}_j$ be the mean of the outcome in group $j$ and $\widehat{\mu}_{cj}$ the mean of all patients who did not drop out. The TM difference, $\beta_t$, is estimated by taking the difference of the observed 50% upper fraction means: $\widehat{\beta}_t = \widehat{\mu}_{t1} - \widehat{\mu}_{t0}$.

In Figure S1A, we observe the full data and in Figure S1B, we have dropout in the placebo group in the lower end of the distribution. For both scenarios, the TM estimator is unbiased, with the true treatment effect ($\beta = 9$) contained within the 95% confidence interval (A: $\widehat{\beta}_t = 9.00$; 95% CI: 8.95,9.04; B: $\widehat{\beta}_t = 9.05$; 95% CI: 8.99,9.09). In contrast, the CCA estimate, calculated from observed means, $\widehat{\mu}_0$ and $\widehat{\mu}_1$, is biased downwards in the presence of worst value dropout ($\widehat{\beta}_c = \widehat{\mu}_0 - \widehat{\mu}_1 = 8.34$; 95% CI: 8.29,8.40).

Figure S1C illustrates a dropout scenario where the location shift assumption is violated, with the placebo group SD exceeding the treatment group SD. As a consequence of the dropout, the observed mean, $\mu_0$, is now too high and the increased placebo SD inflates the upper fraction mean, $\widehat{\mu}_{t0}$, resulting in an underestimation of the treatment effect for the CCA estimate ($\widehat{\beta}_c = 7.93$; 95% CI: 7.87,8.00) and the TM estimate ($\widehat{\beta}_t = 8.16$; 95% CI: 8.09,8.22).

In Figure S1D, we consider equal treatment group SDs, but now the dropout is no longer restricted to the lower half of the distribution, and consequently the strong missing not at random (MNAR) assumption violated. Again, both estimates are biased. The CCA estimate underestimates $\beta$ ($\widehat{\beta}_c = 8.87$; 95% CI: 8.81,8.93), if less so than in Figure S1C, as the dropout is now more evenly spread across the distribution and closer to random dropout. The TM estimate, in contrast, is inflated ($\widehat{\beta}_t = 9.60$; 95% CI: 9.55,9.65).

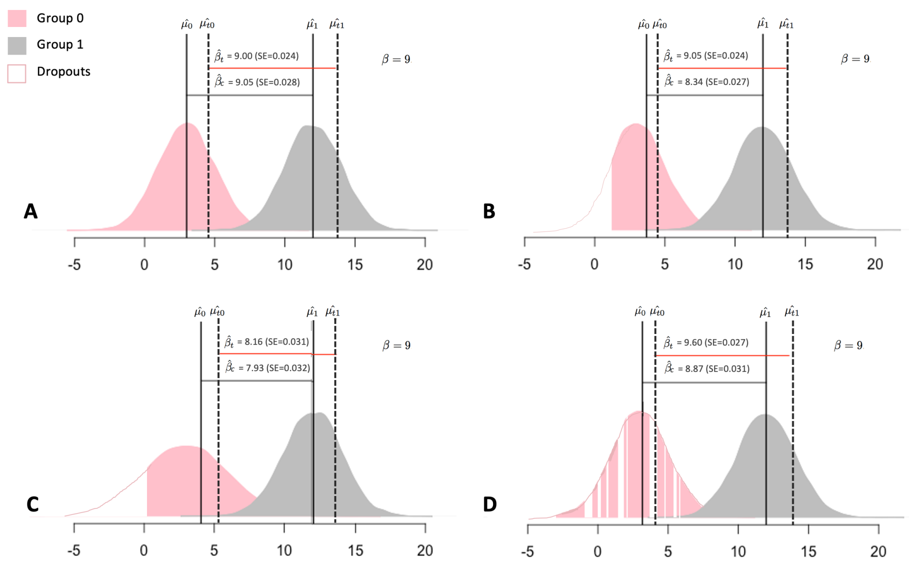

**Figure S1:** Trimmed means (TM) and complete case analysis (CCA) estimators across four scenarios with varying dropout patterns and treatment arm distributions. A) Equal SDs for the placebo (pink) and treatment group (grey) in the absence of dropout. B) Equal treatment group SDs with 20% lower value dropout in the placebo group. C) Unequal treatment group SDs with a greater placebo group SD and 20% lower value placebo group dropout. D) Equal treatment group SDs with 20% dropout ranging across 90% of the placebo group distribution.

### B Location shift assumption bias for a given trimming fraction, $p$, as a function of full sample SDs

Let us consider the location shift assumption and its associated bias $B_{tLS}$:

$$B_{tLS} = \mathbb{E}[\widehat{\beta}_t - \beta] = \mathbb{E}[\beta_t - \beta]. \tag{B1}$$

For normally distributed outcomes, the TM, $\mu_{tj}$, for a given treatment group $j$, can be expressed in terms of a truncated normal distribution. We define the truncated group mean, $\mu_{tj}$, for a distribution with overall mean and SD, $\mu_j$ and $\sigma_j$, with left- and right-truncation at quantiles $p$ and $p_1$, respectively:

$$
\begin{aligned}
\mu_{tj} &= \mu_j - \sigma_j \frac{\phi\big(\Phi^{-1}(p_1)\big) - \phi\big(\Phi^{-1}(p)\big)}{\Phi\big(\Phi^{-1}(p_1)\big) - \Phi^{-1}(\Phi(p))} \\
&= \mu_j - \sigma_j \frac{\phi\big(\Phi^{-1}(p_1)\big) - \phi\big(\Phi^{-1}(p)\big)}{p_1 - p}.
\end{aligned} \tag{B2}
$$

Under the assumption of worse value dropout, we define a mean left-truncated at $p$ ($p_1 = 1$):

$$
\mu_{tj} = \mu_j + \sigma_j \frac{\phi\big(\Phi^{-1}(p)\big)}{1-p}, \tag{B3}
$$

Then, the trimmed population mean difference for a given trimming fraction, $p$, is given by

$$
\beta_t = \mu_{t1} - \mu_{t0} = \left(\mu_1 + \sigma_1 \frac{\phi\big(\Phi^{-1}(p)\big)}{1-p}\right) - \left(\mu_0 + \sigma_0 \frac{\phi\big(\Phi^{-1}(p)\big)}{1-p}\right), \tag{B4}
$$

We define the bias, $B_{tLS}$, with respect to the population mean difference, $\beta = \mu_1 - \mu_0$, for a given trimming fraction, $p$:

$$
B_{tLS} = (\sigma_1 - \sigma_0) \frac{\phi\big(\Phi^{-1}(p)\big)}{1-p}. \tag{B5}
$$

#### C Location shift assumption bias, as a function of trimmed fraction SDs, for a given trimming fraction, $p$, and for 50% trimming ($p = 0.5$)

We now define the TM estimator bias, $B_{tLS}$, in terms of trimmed fraction SDs, for the special case of 50% trimming and for a given trimming fraction, $p$. Consider the general formula for the variance, $\sigma_{tj}^2$, of a truncated normal distribution, with left-truncation at $p$ and right-truncation at $p_1$:

$$\sigma_{tj}^2 = \sigma_j^2 \Big[1 - \frac{\Phi^{-1}(p_1)\phi\big(\Phi^{-1}(p_1)\big) - \Phi^{-1}(p)\phi\big(\Phi^{-1}(p)\big)}{\Phi\big(\Phi^{-1}(p_1)\big) - \Phi\big(\Phi^{-1}(p)\big)} - \Big(\frac{\phi\big(\Phi^{-1}(p_1)\big) - \phi\big(\Phi^{-1}(p)\big)}{\Phi\big(\Phi^{-1}(p_1)\big) - \Phi\big(\Phi^{-1}(p)\big)}\Big)^2\Big]$$
$$= \sigma_j^2 \Big[1 - \frac{\Phi^{-1}(p_1)\phi\big(\Phi^{-1}(p_1)\big) - \Phi^{-1}(p)\phi\big(\Phi^{-1}(p)\big)}{p_1 - p} - \Big(\frac{\phi\big(\Phi^{-1}(p_1)\big) - \phi\big(\Phi^{-1}(p)\big)}{p_1 - p}\Big)^2\Big]. \tag{C1}$$

Then, for a left-truncated distribution ($p_1 = 1$), we have

$$\sigma_{tj}^2 = \sigma_j^2 \Big[1 - \frac{-\Phi^{-1}(p)\phi\big(\Phi^{-1}(p)\big)}{1 - p} - \Big(\frac{-\phi\big(\Phi^{-1}(p)\big)}{1 - p}\Big)^2\Big], \tag{C2}$$

with for 50% trimming

$$\sigma_{tj}^2 = \sigma_j^2 \Big[1 - \Big(\frac{\phi\big(\Phi^{-1}(0.5)\big)}{0.5}\Big)^2\Big] = \sigma_j^2 \Big[1 - \Big(\sqrt{\frac{2}{\pi}}\Big)^2\Big]. \tag{C3}$$

Rearranging (C3) to $\sigma_j^2 = \sigma_{tj}^2/(1 - 2/\pi)$ and substituting in (B5), with $p = 0.5$, gives the bias, $B_{tLS}$, as a function of the 50% trimmed fraction SDs:

$$B_{tLS} = (\sigma_{t1} - \sigma_{t0})\sqrt{\frac{2}{\pi - 2}}. \tag{C4}$$

Equivalently, we can obtain a more generalized bias expression for a given trimming fraction, $p$, by rearranging (C2). Let $Z_p$ denote the quantile of $p$, with $Z_p = \Phi^{-1}(p)$, giving:

$$\sigma_j^2 = \sigma_{tj}^2 \Big/ \Big(1 - \frac{-Z_p\phi(Z_p))}{1 - p} - \Big(\frac{-\phi(Z_p)}{1 - p}\Big)^2\Big), \tag{C5}$$

Then, by substituting (C5) in (B5), we obtain the location shift assumption bias for a general trimming fraction, $p$.

$$B_{tLS} = \Big[\sqrt{\sigma_{t1}^2 \Big/ \Big(1 - \frac{-Z_p\phi(Z_p))}{1 - p} - \Big(\frac{-\phi(Z_p)}{1 - p}\Big)^2\Big)} - \sqrt{\sigma_{t0}^2 \Big/ \Big(1 - \frac{-Z_p\phi(Z_p))}{1 - p} - \Big(\frac{-\phi(Z_p)}{1 - p}\Big)^2\Big]\Big)}\Big]\frac{\phi\big(\Phi^{-1}(p)\big)}{1 - p}. \tag{C6}$$

### D Strong MNAR assumption bias for dropout in the placebo group, for 50% trimming and for a given trimming fraction $p$

Let us consider the bias, $B_{tSM}$, resulting from the strong MNAR assumption violation, supposing that the location shift assumption is satisfied ($\beta_t = \beta$):

$$B_{tSM} = \mathbb{E}[\widehat{\beta}_t - \beta_t] \tag{D1}$$

In Section 2.4 of the main text, we defined the population parameter, $\beta_{td}$, giving the population TM difference for a scenario with a normally distributed treatment group, and a placebo group that is no longer normally distributed due to dropout. Then $\widehat{\beta}_t$ unbiasedly estimates $\beta_{td}$, and with $\beta_t = \mu_{t1} - \mu_{t0}$ and $\beta_{td} = \mu_{t1} - \mu_{td0}$, we write the bias (D1) as

$$B_{tSM} = \mathbb{E}[\beta_{td} - \beta_t] = \mu_{t0} - \mu_{td0}. \tag{D2}$$

Consider a normal distribution with lower bound, $\Phi^{-1}(0)$ and upper bound, $\Phi^{-1}(1)$, and let $f$ denote the fraction of this distribution, so that $f_{0,1} = 1$. Then, let $f_{0,0.5}$ denote the fraction of the distribution that is trimmed away under 50% trimming, $f_{0.5,1}$ the trimmed fraction used for estimation, $f_{0,c}$ the fraction affected by dropout, and $f_{0.5,c}$ the fraction of the distribution for which the trimmed fraction, $f_{0.5,1}$, is affected by dropout. We write the trimmed mean in the absence of dropout as the mean of the truncated distribution corresponding to the fraction, $f_{0.5,1}$: $\mu_t = \mu_{0.5,1}$. In the presence of dropout, the fraction, $f_{0.5,1}$, no longer contains a sufficient number of observations to make up the trimmed fraction, and lower values, originally contained in the fraction, $f_{0,0.5}$, are introduced, resulting in a downwards shift of the lower bound, the trimmed fraction now given by $f_{b,0.5}$, with $b < 0.5$. The trimmed mean in presence of dropout is then given by $\mu_{td} = \mu_{b,1}$.
The shift from 0.5 to $b$ is a function of the dropout proportion, $p_d$, the dropout spread, $f_{0,c}$, the trimming proportion, $f_{0,0.5} = p$, and $f_{0.5,c} = f_{0,c} - p$, with

$$f_{b,c} = \frac{f_{0,c} f_{0.5,c}}{(f_{0,c} - p_d)}, \tag{D3}$$

and

$$b = f_{0,c} - f_{b,c}. \tag{D4}$$

Let us write the TMs as the weighted sum of partial means, and write the trimmed mean in the absence of dropout as $\mu_t = \mu_{0.5,1} = \frac{f_{0.5,c}}{f_{0.5,1}}\mu_{0.5,c} + \frac{f_{c,1}}{f_{0.5,1}}\mu_{c,1}$ and in the presence of dropout as $\mu_{td} = \mu_{b,1} = \frac{f_{0.5,c}}{f_{0.5,1}}\mu_{bc} + \frac{f_{c,1}}{f_{0.5,1}}\mu_{c,1}$. Then, $\mu_{c,1}$ is the mean of a left-truncated normal distribution, and both $\mu_{0.5,1}$ and $\mu_{b,1}$ are the means of bi-truncated normal distributions, with the latter the mean of a

truncated normal only under the assumption of homogeneous dropout in the fraction $f_{bc}$.
We write the bias, $B_{tSM}$ (D2), in terms of weighted partial means:

$$\begin{aligned} B_{tSM} &= \mu_{t0} - \mu_{td0} \\ &= \mu_{0.5,1} - \mu_{b,1} \\ &= \left(\frac{f_{0.5,c}}{f_{0.5,1}}\mu_{0.5,c} + \frac{f_{c,1}}{f_{0.5,1}}\mu_{cd}\right) - \left(\frac{f_{0.5,c}}{f_{0.5,1}}\mu_{bc} + \frac{f_{c,1}}{f_{0.5,1}}\mu_{c,1}\right) \\ &= \frac{f_{0.5,c}}{f_{0.5,1}}\left(\mu_{0.5,c} - \mu_{bc}\right), \end{aligned} \tag{D5}$$

and define $\mu_{bc}$ and $\mu_{0.5,c}$ in terms of a truncated normal distribution, assuming without loss of generality a zero-centered distribution, with $\mu_{bc}$, for example, given by

$$\mu_{bc} = \mu_0 - \sigma_0 \frac{\phi\big(\Phi^{-1}(c)\big) - \phi\big(\Phi^{-1}(b)\big)}{c - b} = \mu_0 - \sigma_0 Q_{bc}, \tag{D6}$$

We write the bias (D5) as a function of the size of the trimmed fraction ($f_{0.5,1}$), the fraction affected by dropout ($f_{0.5,c}$) and the SD of the affected group. Then, for dropout in the placebo group:

$$B_{tSM} = -\frac{f_{0.5,c}}{f_{0.5,1}}\sigma_0(Q_{0.5,c} - Q_{bc}). \tag{D7}$$

Equation (D7) gives the bias on violation of the strong MNAR assumption under 50% trimming. For a generalized trimming fraction $p$ we denote the trimmed fraction $f_{p,1}$, and calculate the shift from $p$ to $b$ with

$$b = f_{0,c} - f_{b,c} = f_{0,c} - \frac{f_{0,c} f_{p,c}}{f_{0,c} - p_d}. \tag{D8}$$

The bias is then given by

$$B_{tSM} = -\frac{f_{p,c}}{f_{p,1}}\sigma_0(Q_{p,c} - Q_{bc}). \tag{D9}$$

### E Strong MNAR assumption bias for dropout in both treatment groups

In Section 2.4 of the main text and in Section D of the appendix, we defined the population parameter, $\beta_{td}$, giving the population TM difference for a scenario with a normally distributed treatment group, and a placebo group that is no longer normally distributed due to dropout. Here, let us write $\beta_{td}$ for the population TM difference given strong MNAR assumption violation in both groups, and define the bias as

$$B_{tSM} = \mathbb{E}[\beta_{td} - \beta_t], \tag{E1}$$

with $\beta_t = \mu_{t1} - \mu_{t0}$, and $\beta_{td} = \mu_{td1} - \mu_{td0}$. As dropout is present in both groups, we write:

$$B_{tSM} = (\mu_{td1} - \mu_{t1}) + (\mu_{t0} - \mu_{td0}) = B_{tSM1} + B_{tSM0}, \tag{E2}$$

distinguishing between a treatment group component and placebo group component, with the former given by $B_{tSM1} = \mu_{td1} - \mu_{t1}$ and the latter by $B_{tSM0} = \mu_{t0} - \mu_{td0}$. In Section 2.4 of the main text and Sections D and E of the appendix, we defined the latter in terms of fractions of a truncated normal distribution:

$$B_{tSM0} = \frac{f_{0.5,c(0)}}{f_{0.5,1}} \left(\mu_{0.5,c(0)} - \mu_{bc(0)}\right), \tag{E3}$$

where $f_{0.5,1}$ denotes the trimmed fraction, $p$, $f_{0.5,c(0)}$ the fraction of the placebo distribution for which the trimmed fraction, $f_{0.5,1}$, is affected by dropout; $\mu_{0.5,c(0)}$ the mean for the fraction, $f_{0.5,c(0)}$, in absence of dropout; and $\mu_{bc(0)}$ the mean in the presence of dropout. The fraction bound, $b_{(0)}$, is calculated from (D3) and (D4) (Section D).

We define the fraction means, $\mu_{0.5,c(0)}$ and $\mu_{bc(0)}$, in terms of truncated normal distributions, as in (D6) (Section D), and write the placebo group strong MNAR assumption bias component as

$$B_{tSM0} = -\frac{f_{0.5,c(0)}}{f_{0.5,1}} \sigma_0 (Q_{0.5,c(0)} - Q_{bc(0)}), \tag{E4}$$

and define the equivalent for the treatment group:

$$B_{tSM1} = -\frac{f_{0.5,c(1)}}{f_{0.5,1}} \sigma_1 (Q_{0.5,c(1)} - Q_{bc(1)}). \tag{E5}$$

The total bias, $B_{tSM}$ (E2), is then given by the sum of (E4) and (E5):

$$\begin{aligned} B_{tSM} = B_{tSM0} + B_{tSM1} = -\frac{f_{0.5,c(0)}}{f_{0.5,1}} \sigma_0 (Q_{0.5,c(0)} - Q_{bc(0)}) - \\ \frac{f_{0.5,c(1)}}{f_{0.5,1}} \sigma_1 (Q_{0.5,c(1)} - Q_{bc(1)}). \end{aligned} \tag{E6}$$

For a given group $j$, in the absence of dropout, $Q_{bc(j)} = Q_{0.5,c(j)}$, and the bias reduces to zero. In the event of dropout in the trimmed fraction, $Q_{bc(j)} <$

$Q_{0.5,c(j)}$, which results in a positive bias component, $B_{tSM0}$ (E4), for the placebo group, and a negative bias component, $B_{tSM1}$ (E5), for the treatment group. For equal dropout proportions across the two groups ($p_{d1} = p_{d0}$) and equal dropout spreads, $Q_{bc(1)} = Q_{bc(0)}$ and $Q_{0.5,c(1)} = Q_{0.5,c(0)}$. Then, given equal SDs ($\sigma_1 = \sigma_0$), the bias components are equally sized, but in opposite directions, and the total bias, $B_{tSM}$ (E6), reduces to zero.

### F A limited upper bound for the strong MNAR assumption bias

Supplementary Figure S2 illustrates the bias that occurs on violation of the strong MNAR assumption in the placebo group, for 20% dropout and under 50% trimming. Figure S2A shows the mean observed bias across $S = 1000$ simulations of sample size $n = 1000$, for a range of dropout spreads and distributions. Figure S2B illustrates the notation used for the fractions, $f$, of the distribution.

For a given dropout spread, $f_{0,c}$, the bias is plotted against the proportion of dropout present in the part of the trimmed fraction that is affected by dropout, $f_{0.5,c}$. The proportion of dropout in the affected fraction ranges from $p_{d(0.5,c)} = 0$, with no dropout in the affected fraction and by extent the 50% trimmed fraction ($f_{0.5,1}$), to $p_{d(0.5,c)} = 1$, when dropout is spread homogeneously across the entirety of $f_{0,c}$. For example, $p_{d(0.5,c)} = 0.6$ for a dropout spread $f_{0,c} = 0.7$, indicates that only 60% of the dropout remains in the affected fraction, $f_{0.5,c} = 0.2$, when compared to the scenario of entirely homogeneous dropout across $f_{0,c} = 0.7$, with the missing 40% now allocated to the fraction that is trimmed away ($f_{0,0.5}$). We observe that the TM estimator is increasingly biased the larger the dropout spread, $f_{0,c}$, and the more homogeneous the dropout, with the observed bias (solid line) consistently lower than the calculated limit (dotted line) for all $p_{d(0.5,c)} < 1$ and equal to it for $p_{d(0.5,c)} = 1$.

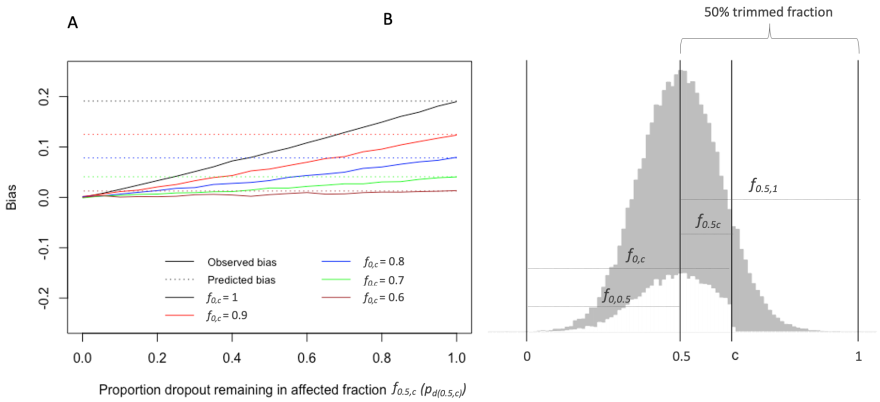

**Figure S2:** Observed and calculated bias on violating the strong MNAR assumption. A) Shown is the mean observed bias across $S = 1000$ simulations of sample size $n = 1000$, with equal treatment group SDs of $\sigma_0 = \sigma_1 = 1$ and 20% dropout in the placebo group. The bias is shown for dropout spreads ($f_{ac}$) of 60 to 100%, across varying dropout distributions, from no dropout in the affected fraction, $f_{0.5,c}$ ($p_{d(0.5,c)} = 0$), to entirely homogeneous dropout across the entirety of $f_{0,c}$ ($p_{d(0.5,c)} = 1$). The bias calculated from (E3, equation 12 in the main text), under the assumption of homogeneity, is shown alongside (dotted line). B) Distributional fractions, $f$.

### G CCA bias under homogeneous dropout in one or both treatment groups

Let us consider the bias, $B_c$, resulting from violation of the random dropout assumption. Previously, in Section 2.4 of the main text, we defined this bias for dropout restricted to the placebo group. Here, we derive expressions for $B_c$ under dropout in both groups:

$$\begin{aligned} B_c &= \mathbb{E}[\beta_c - \beta] \\ &= (\mu_{c1} - \mu_{c0}) - (\mu_1 - \mu_0) \\ &= (\mu_{c1} - \mu_1) + (\mu_0 - \mu_{c0}) \\ &= B_{c1} + B_{c0}, \end{aligned} \tag{G1}$$

with the treatment group bias component given by $B_{c1}$, and placebo group bias component given by $B_{c0}$.

For a given group $j$, we define the full sample mean, $\mu_j$ (G2), and complete case mean, $\mu_{cj}$, (G3) as sums of weighted partial means, with

$$\mu_j = f_{0,c}\mu_{0,c} + (1 - f_{0,c})\mu_{c,1}, \tag{G2}$$

and

$$\mu_{cj} = \frac{(f_{0,c} - p_d)}{(1 - p_d)}\mu_{0,c} + \frac{(1 - f_{0,c})}{(1 - p_d)}\mu_{c,1}, \tag{G3}$$

with $f_{ac}$ denoting the fraction of the distribution affected by dropout, for a distribution bounded by $a = 0$ and $d = 1$. With (G2) and (G3), we define the placebo group bias component $B_{c0} = \mu_0 - \mu_{c0}$:

$$B_{c0} = \left(f_{0,c(0)} - \frac{(f_{0,c(0)} - p_{d0})}{(1 - p_{d0})}\right)\mu_{0,c(0)} + \left((1 - f_{0,c(0)}) - \frac{(1 - f_{0,c(0)})}{(1 - p_{d0})}\right)\mu_{c,1(0)}, \tag{G4}$$

with $f_{0,c(0)}$ the dropout spread, $p_{d0}$ the dropout proportion, $\mu_{0,c(0)}$ the mean of the distribution fraction affected by dropout and $\mu_{c,1(0)}$ the mean of unaffected fraction. We write the partial means in terms of a truncated normal distribution, assuming without loss of generality a zero-centered distribution, e.g., for $\mu_{c,1}$ we write

$$\mu_{c,1} = -\sigma\frac{\phi\big(\Phi^{-1}(1)\big) - \phi\big(\Phi^{-1}(c)\big)}{1 - c}. \tag{G5}$$

With (G4) and (G5), we define the placebo group bias component fully:

$$\begin{aligned} B_{c0} = &-\sigma_0\Bigg[\left(f_{0,c(0)} - \frac{(f_{0,c(0)} - p_{d0})}{(1 - p_{d0})}\right)\frac{\phi\big(\Phi^{-1}(c_0))\big)}{c_0} + \\ &\left((1 - f_{0,c(0)}) - \frac{(1 - f_{0,c(0)})}{(1 - p_{d0})}\right)\frac{-\phi\big(\Phi^{-1}(c_0)\big)}{1 - c_0}\Bigg], \end{aligned} \tag{G6}$$

and, equivalently, for the treatment group bias component, $B_{c1} = \mu_{c1} - \mu_1$, we write:

$$\begin{aligned} B_{c1} = &-\sigma_1\Bigg[\left(\frac{(f_{0,c(1)} - p_{d1})}{(1 - p_{d1})} - f_{0,c(1)}\right)\frac{\phi\big(\Phi^{-1}(c_1))\big)}{c_1} + \\ &\left(\frac{(1 - f_{0,c(1)})}{(1 - p_{d1})} - (1 - f_{0,c(1)})\right)\frac{-\phi\big(\Phi^{-1}(c_1)\big)}{1 - c_1}\Bigg]. \end{aligned} \tag{G7}$$

Given non-random dropout ($p_{dj} > 0$, $f_{ac(j)} < 1$), $B_{c0}$ will be negative, indicating underestimation of the treatment effect as a consequence of overestimation of complete case placebo mean, $\mu_{c0}$, with the converse true for the treatment group bias component, $B_{c1}$. The total bias is then given by the sum of (G6) and (G7), where, under equal dropout proportions ($p_{d1} = p_{d0}$), equal dropout spreads ($f_{ac(1)} = f_{ac(0)}$) and equal SDs ($\sigma_1 = \sigma_0$), the bias components will cancel each other out. In the absence of dropout in either group ($p_{d1} = p_{d0} = 0$) or homogeneous dropout in both groups ($f_{ac(1)} = f_{ac(0)} = 1$), both bias components will reduce to zero.

#### H CCA bias for dropout restricted to the lowest part of the distribution

Here, we derive the CCA bias under dropout restricted to the lowest part of the distribution, for the case of dropout in the placebo group. We define $B_{c0}$, as in (G6):

$$B_{c0} = -\sigma_0 \Bigg[ \left( f_{0,c(0)} - \frac{(f_{0,c(0)} - p_{d0})}{(1 - p_{d0})} \right) \frac{\phi\big(\Phi^{-1}(c0)\big)}{c0} + \left( (1 - f_{0,c(0)}) - \frac{(1 - f_{0,c(0)})}{(1 - p_{d0})} \right) \frac{-\phi\big(\Phi^{-1}(c0)\big)}{1 - c0} \Bigg]. \tag{H1}$$

with $f_{0,c(0)}$ the dropout spread, $p_{d0}$ the dropout proportion, and $c_0$ the upper bound of $f_{0,c(0)}$, for a distribution bounded by $\Phi^{-1}(0)$ and $\Phi^{-1}(1)$.

For the special case of dropout restricted to the very lower end of the distribution, $f_{0,c(0)} = p_{d0} = c_0$, and (H1) reduces to

$$\begin{aligned} B_{c0} &= -\sigma_0 \left[ p_{d0} \frac{\phi\big(\Phi^{-1}(p_{d0})\big)}{p_{d0}} - p_{d0} \frac{-\phi\big(\Phi^{-1}(p_{d0})\big)}{1 - p_{d0}} \right] \\ &= -\sigma_0 \left[ \frac{\phi\big(\Phi^{-1}(p_{d0})\big)(1 - p_{d0})}{1 - p_{d0}} - \frac{-\phi\big(\Phi^{-1}(p_{d0})\big) p_{d0}}{1 - p_{d0}} \right] \\ &= -\sigma_0 \frac{\phi\big(\Phi^{-1}(p_{d0})\big)}{1 - p_{d0}}. \end{aligned} \tag{H2}$$

Dropout of lower values leads to an overestimation of the complete case mean, $\mu_{c0}$. The placebo CCA bias component, $B_{c0}$, is negative, and reflects the underestimation of the treatment effect, resulting from the overestimated $\mu_{c0}$. The same bias quantity can be calculated for dropout in the treatment group, by substituting (G7) for (H1) and solving from there, giving:

$$B_{c1} = \sigma_1 \frac{\phi\big(\Phi^{-1}(p_{d1})\big)}{1 - p_{d1}}. \tag{H3}$$

For dropout in the treatment group, the bias, $B_{c1}$, will be positive, reflecting the overestimation of the treatment effect as a consequence of the overestimated treatment group complete case mean, $\mu_{c1}$.

A similar principle extends to dropout at the upper end of the distribution. The bias, owing to the symmetry of the normal distribution, is equal in size and opposite to the bias calculated under the assumption of lowest value dropout (e.g., if for lowest value dropout, $B_{C1} = x$, then for highest value dropout, $B_{C1} = -x$). For a given treatment group, and dropout restricted to either the very lowest or highest values, the group-specific bias component, $B_{Cj}$, is the maximum bias and given by

$$\mid B_{Cj} \mid = \frac{\sigma_{0j}}{(1 - p_{dj})} \phi\big(\Phi^{-1}(p_{dj})\big). \tag{H4}$$

### I Maximum bias of the TM estimator in relation to Copas' and Jackson's bias limit

Copas and Jackson [7] defined a maximum bias for selection models, and proved that this bias limit is achieved when the selection model takes the form of a threshold function, with the selection probability, $p_s$, of the outcome, $y$, going from 0 to 1 at threshold, $t$:

$$p_s = \begin{cases} 1 & \text{if } y \geq t \\ 0 & \text{if } y < t. \end{cases} \tag{I1}$$

For a given selection probability, $p_s$, they define the maximum bias, $B_{\max}$, as a function of the full sample SD, $\sigma$:

$$|B_{\max}| = \frac{\sigma}{p_s} \phi\big(\Phi^{-1}(p)\big). \tag{I2}$$

The maximum bias for a given group under the CCA estimator (H4) is obtained under either strict lowest value dropout or strict highest value dropout, and is equivalent to Copas and Jackson's maximum bias. Here, we extend this formula and derive expressions for the maximum bias of the TM estimator.

Let us consider the TM estimator in context of a threshold selection model. In order for the strong MNAR assumption to be satisfied, it is sufficient that all dropouts are contained in the fraction, $p$, that is trimmed away, or, equivalently, that the trimmed fraction used for estimation $(1 - p)$ is completely observed. When trimming under assumption of lower value dropout, all dropout values should be lower than the distribution quantile corresponding to $p$:

$$p_s = \begin{cases} [0,1] & \text{if } y < \mu + \sigma\Phi^{-1}(p) \\ 1 & \text{if } y \geq \mu + \sigma\Phi^{-1}(p). \end{cases} \tag{I3}$$

The bias resulting from the violation of (I3) is a function of the overall selection probability, $p_s$, and of the trimmed fraction, $1-p$. Let us define the maximum possible TM estimator bias, which, under the worst case scenario given in the case of lower value trimming, occurs under highest value dropout. For $p_{dj}$ highest value dropout, all observations exceeding the quantile corresponding to $1 - p_{dj}$ are unobserved:

$$p_s = \begin{cases} 0 & \text{if } y \geq \mu_j + \sigma_j\Phi^{-1}(1-p_{dj}) \\ 1 & \text{if } y < \mu_j + \sigma_j\Phi^{-1}(1-p_{dj}). \end{cases} \tag{I4}$$

The TM estimator bias is then given by the difference between the mean, $\mu_{tdj}$, obtained under the selection model of (I4), but the assumption of (I3), and the mean, $\mu_{tj}$, obtained under the selection model of (I3). Then, for $p_d$ highest value dropout, and lower value trimming of fraction, $p$, $\mu_{tdj}$ is calculated over the $1-p$ trimmed fraction, which is given from $(1-p_{dj})-(1-p) = p_{sj}-(1-p)$ to $(1-p_{dj}) = p_{sj}$. We define the maximum bias, $B_{tSM0\max}$, under highest value dropout in the placebo group and lower value $p$ trimming:

$$B_{tSM0\max} = \mu_{t0} - \mu_{td0}, \tag{I5}$$

with

$$\mu_{t0} = \mu_0 + \sigma_0 \frac{\phi\big(\Phi^{-1}(p)\big)}{1-p}, \tag{I6}$$

and

$$\mu_{td0} = \mu_0 - \sigma_0 \frac{\phi\big(\Phi^{-1}(p_{s0})\big) - \phi\big(\Phi^{-1}(p_{s0}-(1-p))\big)}{1-p}, \tag{I7}$$

with $p_{s0} = 1 - p_{d0}$.

The maximum bias is then given by

$$\begin{aligned} B_{tSM0\max} =& \frac{\sigma_0}{1-p}\Big[\phi\big(\Phi^{-1}(p)\big) - -\phi\big(\Phi^{-1}(p_{s0})\big) - \\ & \phi\big(\Phi^{-1}(p_{s0}-(1-p))\big)\Big]. \end{aligned} \tag{I8}$$

Equation (I8) extends Copas' and Jackson's original formula (I2) to the TM estimator, giving the bias on maximal violation of the strong MNAR assumption. To account for potential violations of the location shift assumption bias, we add the previously defined $B_{tLS}$ bias component (B5):

$$
\begin{aligned}
B_{t0\max} =& \frac{\sigma_0}{1-p}\Big[\phi\big(\Phi^{-1}(p)\big) - -\phi\big(\Phi^{-1}(p_{s0})\big) - \\
& \phi\big(\Phi^{-1}(p_{s0}-(1-p))\big)\Big] + \\
& \Big(\sigma_1 - \sigma_0\Big)\frac{\phi\big(\Phi^{-1}(p)\big)}{1-p}.
\end{aligned} \tag{I9}
$$

Equally, we can derive the maximum bias for dropout in the treatment group, with $B_{tSM1\max}$ given by

$$
B_{tSM1\max} = \mu_{td1} - \mu_{t1}. \tag{I10}
$$

Then, with $\mu_{t1}$ and $\mu_{td1}$ defined analogously to $\mu_{t0}$ (I6) and $\mu_{td0}$ (I7),

$$
\begin{aligned}
B_{tSM1\max} = \frac{\sigma_1}{1-p}\Big[ & -\Big(\phi\big(\Phi^{-1}(p_{s1})\big) - \phi\big(\Phi^{-1}(p_{s1}-(1-p))\big)\Big) - \\
& \phi\big(\Phi^{-1}(p)\big)\Big],
\end{aligned} \tag{I11}
$$

and

$$
\begin{aligned}
B_{t1\max} = \frac{\sigma_1}{1-p}\Big[ & -\Big(\phi\big(\Phi^{-1}(p_{s1})\big) - \phi\big(\Phi^{-1}(p_{s1}-(1-p))\big)\Big) \\
& - \phi\big(\Phi^{-1}(p)\big)\Big] + \Big(\sigma_1 - \sigma_0\Big)\frac{\phi\big(\Phi^{-1}(p)\big)}{1-p}.
\end{aligned} \tag{I12}
$$

#### J Simulation illustration: CCA and TM estimator bias for highest value placebo group dropout

Consider a clinical trial on $n = 1000$ subjects randomized to an intervention ($j = 1$) and placebo ($j = 0$) arm, with $n_1 = n_0 = 500$, normally distributed outcomes $y$, a true treatment effect $\beta = 0.5$ and 20% strict highest value dropout in the placebo group. Supplementary Table S1 gives a simulated example ($S = 1000$) for three different placebo group SDs, $\sigma_0$, with the treatment group SD, $\sigma_1$, held constant. Mean treatment effect estimates are obtained for the CCA, $\widehat{\beta}_c$, and TM estimators, $\widehat{\beta}_t$, for the latter under the assumption of lower value dropout.

**Table S1:** Treatment effects for normally distributed outcomes with treatment and placebo group SDs $\sigma_1 = 1$ and $\sigma_0 = 0.5, 1, 1.5$, true treatment effect $\beta = 0.5$, for 20% highest value placebo group dropout, and 50% lower value trimming. Shown are the mean CCA ($\widehat{\beta}_c$) and TM ($\widehat{\beta}_t$) estimates across $S = 1000$ simulations ($N = 1000$). Also shown are the bias components: the CCA estimator bias, $\widehat{B}_C$; the TM estimator location shift assumption bias, $\widehat{B}_{tLS}$; the TM estimator strong MNAR bias, $\widehat{B}_{tSM}$; the total TM estimator bias, $\widehat{B}_t$.

| $\sigma_0$ | $\widehat{\beta}$ | $\widehat{\beta}_c$ | $\widehat{\beta}_t$ | $\widehat{B}_c$ | $\widehat{B}_{tLS}$ | $\widehat{B}_{tSM}$ | $\widehat{B}_t$ |
|---|---|---|---|---|---|---|---|
| 0.5 | 0.50 | 0.68 | 1.23 | 0.18 | 0.40 | 0.33 | 0.73 |
| 1 | 0.50 | 0.85 | 1.16 | 0.35 | 0.00 | 0.66 | 0.66 |
| 1.5 | 0.50 | 1.03 | 1.10 | 0.53 | -0.40 | 0.99 | 0.59 |

For the CCA estimator, higher value dropout in the placebo group will lead to underestimation of the placebo complete case mean and overestimation of the treatment effect. From Supplementary Table S1, we see that $\widehat{\beta}_c > \widehat{\beta}$ for all placebo group SDs, with the greatest bias for $\sigma_0 = 1.5 > \sigma_1$. Consider the maximum CCA bias, previously defined in (H3). For highest value placebo dropout, we can then write

$$B_{C0\max} = \frac{\sigma_0}{(1-p_d)}\phi\big(\Phi^{-1}(p_d)\big). \tag{J1}$$

Filling in (J1), for $\sigma_0 = 1$ and $p_d = 0.2$, gives

$$B_{C0\max} = \frac{1}{(1-0.2)}\phi\big(\Phi^{-1}(0.2)\big) = 0.35,$$

equalling the mean CCA bias, $\widehat{B}_C = 0.35$, observed for $\sigma_0 = 1$.

For the TM estimator, higher value dropout in the placebo group, given lower value trimming, will also lead to underestimation of the placebo trimmed mean and overestimation of the treatment effect. This effect can either be exacerbated or mitigated by unequal treatment group SDs, with $\sigma_0 < \sigma_1$ giving a positive location shift assumption bias component, $B_{tLS}$, and $\sigma_0 > \sigma_1$ contributing a negative component, $B_{tLS}$.

In Supplementary Table S1, we observe positive bias components, $\widehat{B}_{tSM}$, resulting from strong MNAR assumption violation, with the greatest bias for the largest $\sigma_0$. We observe a positive component, $\widehat{B}_{tLS} = 0.40$ for $\sigma_0 = 0.5 < \sigma_1$, and a negative component, $\widehat{B}_{tLS} = -0.40$ for $\sigma_0 = 1.5 > \sigma_1$, with the total observed TM estimator bias given by $\widehat{B}_t$. Consider the maximum TM estimator bias on violation of the strong MNAR assumption, previously defined in (I8):

$$B_{tSM0\max} = \frac{\sigma_0}{1-p}\Big[\phi\big(\Phi^{-1}(p_s)\big) - \phi\big(\Phi^{-1}(p_s-(1-p))\big) + \phi\big(\Phi^{-1}(1-p)\big)\Big]. \tag{J2}$$

Filling in $p = 0.5$, $p_s = 1 - p_d = 0.8$ and $\sigma_0 = 1$, gives

$$B_{tSM0\max} = \frac{1}{0.5}\left[\phi\big(\Phi^{-1}(0.8)\big) - \phi\big(\Phi^{-1}(0.3)\big) + \phi\big(\Phi^{-1}(0.5)\big)\right] = 0.66,$$

which corresponds to the strong MNAR bias component of the TM estimator, $\widehat{B}_{tSM}$, observed for $\sigma_0 = 1$ in Supplementary Table S1.

### K Adjusted estimator bias under strong MNAR violation in either or both treatment groups

Here, we define the adjusted estimator bias for the following four scenarios: **K.1**) the bias on rescaling the placebo group, given strong MNAR violation in the placebo group;**K.2**) the bias on rescaling the placebo group, given strong MNAR violation in both the placebo and treatment groups; **K.3**) the bias on rescaling the treatment group, given strong MNAR violation in the placebo group; **K.4**) the bias on rescaling the treatment group, given strong MNAR violation in the treatment group and optionally the placebo group. In Section **K.5**, formulae are derived for the mirrored full sample variance, $\sigma^2_{mj}$, for a given group $j$, in the absence and presence of dropout in the mirrored fraction.

#### K.1 Adjusted estimator bias on rescaling the placebo group, given strong MNAR violation in the placebo group

In Section 2.5 of the main text we defined the adjusted TM estimator for placebo group rescaling, and showed that under the strong MNAR assumption, this adjustment gives an unbiased treatment effect estimate when the location shift assumption is violated (equation 20 in the main text). This adjustment is performed for 50% trimming. Under the assumption of normally distributed outcomes, the 50% trimmed fraction will be distributed half-normally, and the SD of a given group can be adjusted by mirroring the fraction and rescaling the resulting distribution, using the full sample SD of the other group. The population adjusted TM estimate, on rescaling the placebo group, is given by

$$\beta_{at0} = \mu_{t1} - \mu_{at0}, \tag{K1}$$

with $\mu_{t1}$ the unadjusted 50% treatment group TM, $\mu_{at0}$ the adjusted 50% placebo group TM:

$$\mu_{at0} = \frac{\mu_{t0} - \mu_0}{\sigma_0/\sigma_1} + \mu_0, \tag{K2}$$

with $\mu_{t0}$ the unadjusted 50% placebo TM:

$$\mu_{t0} = \mu_0 + \sigma_0 \phi\big(\Phi^{-1}(0.5)\big)/0.5. \tag{K3}$$

We now consider this adjustment under strong MNAR assumption violation. The full sample SD ($\sigma_0$) and mean ($\mu_0$) in (K2) remain unobserved and are inferred in the process of mirroring. We denote the SD and mean of the mirrored distribution $\sigma_{m0}$ and $\mu_{m0}$, and write $\mu_{t0}$ as $\mu_{td0}$, which is the 50% placebo trimmed mean under strong MNAR assumption violation. The adjusted trimmed mean under strong MNAR assumption violation can then be written as

$$\mu_{atd0} = \frac{\mu_{td0} - \mu_{m0}}{\sigma_{m0}/\sigma_1} + \mu_{m0}, \tag{K4}$$

with $\sigma_1$ being the full sample SD from the treatment group, which is either observed in absence of treatment group dropout, or extrapolated from the 50% trimmed fraction. The adjusted TM estimate is given by

$$\beta_{at0d} = \mu_{t1} - \mu_{atd0}.$$

We define the adjusted estimator bias, $B_{at0}$, for dropout in the placebo group, and under placebo group rescaling:

$$\begin{aligned} B_{at0} &= \beta_{at0d} - \beta_{at0} \\ &= (\mu_{t1} - \mu_{atd0}) - (\mu_{t1} - \mu_{at0}) \\ &= \mu_{at0} - \mu_{atd0}. \end{aligned} \tag{K5}$$

From (K2) and (K4) we note that $\mu_{at0}$ and $\mu_{atd0}$ will be identical when $\mu_{td0} = \mu_{t0}$, $\sigma_{m0} = \sigma 0$, and $\mu_{m0} = \mu_0$. These equalities are subject to the strong MNAR assumption and the assumption of distributional normality holding. On violation of the former, $\mu_{td0}$ is underestimated, and $\sigma_{m0}$ overestimated, resulting in the underestimation of $\mu_{atd0}$, and a positive bias, $B_{at0}$.

Let us write the mean of a given distributional fraction, $a$ to $b$, as $-\sigma_j Q_{ab}$, with the Q-term the mean of the standard normal for the relevant fraction, expressed in terms of the pdf and quantile functions ($Q_{ab} = \big(\phi\big(\Phi^{-1}(b)\big) - \phi\big(\Phi^{-1}(a)\big)\big)/(b-a)$). From equation 20 in the main text, we observe that $\mu_{at0}$ is the placebo 50% trimmed mean under the treatment SD, $\sigma_1$. Assuming, without loss of generality, a zero-centered distribution, we write $\mu_{at0} = -\sigma_1 Q_{0.5,1}$, and $\mu_{td0} = -\sigma_0(f_1 Q_{bc} + f_2 Q_{c,1})$, where $\mu_{td0}$ is the placebo 50% trimmed mean under violation for the strong MNAR assumption, written as the sum of weighted fraction means, for fractions $f_1 = f_{0.5,c}/f_{0.5,1}$ and $f_2 = f_{c,1}/f_{0.5,1}$, and with $b$ obtained from (D3, D4). Under a zero-centered distribution, no dropout and strict normality, the placebo mean of the mirrored 50% trimmed fraction, $\mu_{0m}$, is given by $\mu_{0m} = \sigma_0 Z_{0.5} = 0$, with $Z_{0.5}$ the 0.5 quantile of the standard normal distribution. Under strong MNAR assumption violation, $b < 0.5$ and $Z_b < 0$.

We define $B_{at0}$ (K5), fully as

$$B_{at0} = -\sigma_1 Q_{0.5,1} - \Big( - \frac{\sigma_1\sigma_0}{\sigma_{m0}}(f_1 Q_{bc} + f_2 Q_{c,1}) - \frac{\sigma_1\sigma_0}{\sigma_{m0}} Z_b + \sigma_0 Z_b \Big), \qquad \text{(K6)}$$

for a distribution with dropout spread, $f_{0,c}$, 50% trimmed fractions $f_{0.5,1}$ and $f_{b,1}$ in absence and presence of dropout, respectively, and with $Z_b$ the $b$ quantile of a standard normal distribution.

We can distinguish three sources of bias as a consequence of strong MNAR assumption violation: underestimation of $\mu_{td0}$ though $Q_{b,1}$, overestimation of the mirrored placebo distribution SD, $\sigma_{0m}$, and the contribution of $Z_b$, which is no longer 0.

Given strong MNAR violation, lower values will be introduced into the 50% trimmed fraction, resulting in a lower 50% trimmed mean ($\mu_{td0} = \sigma_0 Q_{bd}$), and a comparatively larger fraction SD, which in turn results in an inflated mirrored full sample SD. With $\sigma_{0m} > \sigma_0$, $\frac{\sigma_1\sigma_0}{\sigma_{m0}} < \sigma_1$, and $\mu_{atd0}$ is biased further downwards. The remaining component of (K6), $\frac{\sigma_1\sigma_0}{\sigma_{m0}} Z_b + \sigma_0 Z_b$, will result in either a further bias downwards, for $\sigma_0 > \sigma_1$, or will partly mitigate it, for $\sigma_0 < \sigma_1$. Formulae for the mirrored full sample variance $\sigma_{m0}$, in presence and absence of dropout are derived in Section K.5.

#### K.2 Adjusted estimator bias on rescaling the placebo group, given strong MNAR violation in both treatment groups

Given dropout in the placebo and treatment group, we write the bias on rescaling the placebo group as

$$\begin{aligned} B_{at0} &= \beta_{at0} - \beta_t \\ &= (\mu_{td1} - \mu_{atd0}) - (\mu_{t1} - \mu_{at0}) \\ &= (\mu_{at0} - \mu_{atd0}) + (\mu_{td1} - \mu_{t1}) \\ &= (\mu_{at0} - \mu_{atd0}) + B_{tSM1}, \end{aligned} \qquad \text{(K7)}$$

with $B_{tSM1}$ (E5) a positive bias component, resulting from underestimation of the treatment group trimmed mean under strong MNAR assumption violation (Appendix E). Violation of the strong MNAR assumption in the treatment group will mitigate the negative bias resulting from the underestimation of $\mu_{atd0}$ in two ways. First, by introducing a positive bias component to counteract the negative one; secondly, by counteracting the overestimated mirrored placebo group SD, $\sigma_{m0}$, as the SD extrapolated from the 50% trimmed treatment group fraction is now also inflated. Let us denote the extrapolated treatment group SD by $\sigma_{e1}$, and write the total bias as

$$B_{at0} = -\sigma_1 Q_{0.5,1} - \Big( -\frac{\sigma_{e1}\sigma_0}{\sigma_{m0}}(f_1 Q_{bc} + f_2 Q_{c,1}) - \frac{\sigma_{e1}\sigma_0}{\sigma_{m0}} Z_b + \sigma_0 Z_b \Big) + B_{tSM1}, \tag{K8}$$

with $B_{tSM1}$ obtained from (E5), and $\sigma_{e1}$ and $\sigma_{m0}$ calculated as detailed in Section K.5, from (K17), (K19) and (C1).

#### K.3 Adjusted estimator bias on rescaling the treatment group, given strong MNAR violation in the placebo group

Let us now consider the adjusted TM estimate obtained on rescaling the treatment group, under assumption of normality and for 50% trimming. We define $\beta_{at1}$ and $\beta_{atd1}$, with $\beta_{at1} = \mu_{at1} - \mu_{t0}$ an unbiased estimate of $\beta_t$, and $\beta_{atd1} = \mu_{atd1} - \mu_{t0d}$ the adjusted TM estimate under strong MNAR assumption violation in the placebo group. The adjusted estimator bias for dropout in the placebo group, under treatment group rescaling is then given by:

$$\begin{aligned} B_{at1} &= \beta_{atd1} - \beta_{at1} \\ &= (\mu_{atd1} - \mu_{td0}) - (\mu_{at1} - \mu_{t0}) \\ &= (\mu_{atd1} - \mu_{at1}) + (\mu_{td1} - \mu_{t1} \\ &= (\mu_{atd1} - \mu_{at1}) + B_{tSM0}, \end{aligned} \tag{K9}$$

with $B_{tSM0}$ being the bias when violating strong MNAR assumption in placebo group from (E4), and $\mu_{at1}$ and $\mu_{atd1}$ defined analogously to $\mu_{at0}$ (K2) and $\mu_{atd0}$ (K4), with

$$\mu_{at1} = \frac{\mu_{t1} - \mu_1}{\sigma_1/\sigma_0} + \mu_1, \tag{K10}$$

and

$$\mu_{atd1} = \frac{\mu_{td1} - \mu_{m1}}{\sigma_{m1}/\sigma_{e0}} + \mu_{m1}. \tag{K11}$$

As the strong MNAR assumption is satisfied for the treatment group, $\mu_{td1} = \mu_{t1}$, and $\mu_{m1} = \mu_1 = 0$, and $\sigma_{m1} = \sigma_1$, with $\sigma_{e0}$ being the only source of bias affecting the adjusted treatment TM, $\mu_{atd1}$, resulting in a comparatively slight overestimation of the treatment group TM, and a positive bias:

$$\begin{aligned} B_{at1} &= \Big( -\frac{\sigma_{e0}\sigma_1}{\sigma_1} Q_{0.5,1} \Big) - (-\sigma_0 Q_{0.5,1}) + B_{tSM0} \\ &= (\sigma_0 - \sigma_{e0}) Q_{0.5,1} + B_{tSM0}. \end{aligned} \tag{K12}$$

#### K.4 Adjusted estimator bias on rescaling the treatment group, given strong MNAR violation in the treatment group and optionally in the placebo group

Finally, let us consider a scenario where the strong MNAR assumption is violated in the treatment group, rather than the placebo group, and we rescale the treatment group, to obtain $\beta_{at1}$. Then, with (K9), analogous to (K6), we define the adjusted estimator bias, which will be negative and given by:

$$B_{at1} = \Big( -\frac{\sigma_0 \sigma_1}{\sigma_{m1}}(f_1 Q_{bc} + f_2 Q_{c,1}) - \frac{\sigma_0 \sigma_1}{\sigma_{m1}} Z_b + \sigma_1 Z_b \Big) - (-\sigma_1 Q_{0.5.1}), \quad \text{(K13)}$$

and, in the event of strong MNAR violation in the placebo group, by

$$\begin{aligned} B_{at1} = &\Big( -\frac{\sigma_{e0} \sigma_1}{\sigma_{m1}}(f_1 Q_{bc} + f_2 Q_{c,1}) - \frac{\sigma_{e0} \sigma_1}{\sigma_{m1}} Z_b + \sigma_1 Z_b \Big) - \\ &(-\sigma_1 Q_{0.5,1}) + B_{tSM0}. \end{aligned} \quad \text{(K14)}$$

On rescaling the placebo group for this same scenario, with dropout only in the treatment group, the adjusted estimator bias will, analogous to (K12), be given by

$$B_{at0} = (\sigma_1 - \sigma_{e1}) Q_{0.5,1} + B_{tSM1}. \quad \text{(K15)}$$

#### K.5 Obtaining the mirrored full sample SD under strong MNAR assumption violation

As a result of strong MNAR violation, the SD of the mirrored distribution for a given group $j$, $\sigma_{jm}$, is overestimated ($\sigma_{jm} > \sigma_j$). The variance of the trimmed fractions, $f_{0.5,1}$ or $f_{b,1}$, can be written as a function of two non-overlapping sub-fractions, $f_{0.5,c}$ and $f_{c,1}$, using the underlying sample means, variances and fraction sizes:[8]

$$\begin{aligned} \sigma^2_{0.5,1} &= \frac{1}{f_1 + f_2}\Big[ f_1 \sigma^2_{0.5,c} + f_2 \sigma^2_{c,1} + \frac{f_1 f_2}{f_1 + f_2}(\mu_{0.5,c} - \mu_{c,1})^2 \Big] \\ &= f_1 \sigma^2_{0.5,c} + f_2 \sigma^2_{c,1} + f_1 f_2 (\mu_{0.5,c} - \mu_{c,1})^2, \end{aligned} \quad \text{(K16)}$$

and equivalently for the fraction, $f_{b,1}$:

$$\sigma^2_{b,1} = f_1 \sigma^2_{bc} + f_2 \sigma^2_{c,1} + f_1 f_2 (\mu_{bc} - \mu_{c,1})^2, \quad \text{(K17)}$$

with $f_1 = \frac{f_{0.5,c}}{f_{0.5,1}}$, $f_2 = \frac{f_{c,1}}{f_{0.5,1}}$, and the variance of a given distribution fraction given by the variance of a truncated normal distribution, calculated from (C1).

It is clear that the fraction variance under dropout, $\sigma^2_{bd}$, will exceed the variance in absence of dropout, $\sigma^2_{0.5,1}$, as the straightforward consequence of a larger fraction being evaluated for the former ($b > 0.5$). This is equally apparent from equations (K16) and (K17), with $\sigma^2_{bc} > \sigma^2_{0.5,c}$ and $(\mu_{bc} - \mu_{c,1}) > (\mu_{0.5,c} - \mu_{c,1})$.

From the fraction variances, $\sigma^2_{0.5,1}$ and $\sigma^2_{b,1}$, the variance of the mirrored distribution, $\sigma^2_{m0}$, is easily obtained through equation (K16), now applied to the observed fraction, $f_{0.5,1}$, and the mirrored fraction, $f'_{0.5,1}$, with $f_1 = f_2 = 0.5$. In the absence of dropout, we then have:

$$\sigma^2_{mj} = \sigma^2_j = \sigma^2_{0.5,1} + 0.25(\mu_{0.5,1} - \mu'_{0.5,1})^2, \tag{K18}$$

while in the presence of dropout we obtain:

$$\sigma^2_{mj} = \sigma^2_{b,1} + 0.25(\mu_{b,1} - \mu'_{b,1})^2. \tag{K19}$$

The variance of the mirrored distribution in absence of dropout (K18) can also be calculated using normal distribution rules, as previously shown in (C3), with $\sigma^2_j = \frac{\sigma^2_{0.5,1}}{(1-2/\pi)}$.

#### L Companion to Table 1: Examples of bias calculations

Let us consider the simulation reported in Table 1 in the main text, and employ the bias formulae to calculate the CCA estimator bias, $B_c$, the location shift assumption bias, $B_{tLS}$, the strong MNAR assumption bias, $B_{tSM}$, and the adjusted TM estimator bias under placebo group rescaling, $B_{at0}$. For simplicity, let us consider a single scenario from Table 1.b in the main text, in which both assumptions are violated, with 20% dropout ($p_d = 0.2$) spread homogeneously across 75% of the placebo distribution, for a placebo SD, $\sigma_0 = 1.5$, and a treatment group SD, $\sigma_1 = 1$.

##### L.1 CCA bias

Consider the CCA bias on violation of the random dropout assumption in the placebo group, defined in (G6) and equation 14 in the main text. Then, for $\sigma_0 = 1.5$, dropout proportion, $p_d = 0.2$, dropout spread, $f_{0,c} = 0.75$ with upper bound, $c = 0.75$, the bias is given by

$$
\begin{aligned}
B_C = -\sigma_0 \Bigg[ & \left( f_{0,c} - \frac{(f_{0,c} - p_d)}{(1-p_d)} \right) \frac{\phi\big(\Phi^{-1}(c)\big)}{c} + \\
& \left( (1-f_{0,c}) - \frac{(1-f_{0,c})}{(1-p_d)} \right) \frac{-\phi\big(\Phi^{-1}(c)\big)}{1-c} \Bigg] \\
= -1.5 \Bigg[ & \left( 0.75 - \frac{(0.75-0.2)}{(1-0.2)} \right) \frac{\phi\big(\Phi^{-1}(0.75)\big)}{0.75} + \\
& \left( (1-0.75) - \frac{(1-0.75)}{(1-0.2)} \right) \frac{-\phi\big(\Phi^{-1}(0.75)\big)}{1-0.75} \Bigg] \\
= -1.5 * 0.11 = -0.16
\end{aligned}
\tag{L1}
$$

#### L.2 Location shift assumption bias

Consider the location shift assumption bias defined in (B5) and equation 6 in the main text. Then, for $\sigma_1 = 1$, $\sigma_0 = 1.5$ and trimming fraction, $p = 0.5$, the bias is given by

$$
B_{tLS} = (\sigma_1 - \sigma_0) \frac{\phi\big(\Phi^{-1}(p)\big)}{1-p} = (1-1.5) \frac{\phi\big(\Phi^{-1}(0.5)\big)}{0.5} = -0.40, \tag{L2}
$$

#### L.3 Strong MNAR assumption bias

Let us define $b = f_{0,c} - f_{bc}$, with $f_{bc}$ from (D3):

$$
f_{bc} = \frac{f_{0,c(0)} f_{0.5,c(0)}}{(f_{0,c(0)} - p_{d0})} = \frac{0.75 * 0.25}{0.75 - 0.2} = 0.34, \tag{L3}
$$

giving $b = f_{0,c} - f_{bc} = 0.75 - 0.34 = 0.41$. Consider the placebo group component of the strong MNAR assumption bias from (E4) and equation 12 in the main text, with $-\sigma_0 Q_{ab}$ the truncated normal mean for a given fraction $a$ to $b$ as in (D6). The bias is given by

$$\begin{aligned} B_{tSM0} &= -\frac{f_{0.5,c(0)}}{f_{0.5,1}}\sigma_0(Q_{0.5,c(0)} - Q_{bc(0)}) \\ &= \frac{0.25}{0.5} \times -1.5\Big(\frac{\phi\big(\Phi^{-1}(0.75)\big) - \phi\big(\Phi^{-1}(0.5)\big)}{0.75 - 0.5} - \\ &\frac{\phi\big(\Phi^{-1}(0.75)\big) - \phi\big(\Phi^{-1}(0.409)\big)}{0.75 - 0.409}\Big) \\ &= 1.5 \times 0.059 = 0.09. \end{aligned} \tag{L4}$$

#### L.4 Adjusted TM estimator bias

Let us now consider the adjusted estimator bias, for dropout in the placebo group only, and for rescaling the placebo group. The bias (K6) is given by

$$B_{at0} = -\sigma_1 Q_{0.5,1} - \Big(-\frac{\sigma_1\sigma_0}{\sigma_{m0}}(f_1 Q_{bc} + f_2 Q_{c,1}) - \frac{\sigma_1\sigma_0}{\sigma_{m0}} Z_b + \sigma_0 Z_b\Big). \tag{L5}$$

Components $Q_{0.5,1}$ and $Q_{b,c}$ are truncated standard normal distribution means, with $c = 0.75$, and $b = 0.41$, from (L3), giving

$$Q_{bc} = \frac{\phi\big(\Phi^{-1}(0.75)\big) - \phi\big(\Phi^{-1}(0.41)\big)}{(0.75 - 0.41)} = -0.21, \tag{L6}$$

with $Q_{bd} = -0.80$ calculated in the same manner.

Treatment and placebo group SDs are known, with $\sigma_1 = 1$ and $\sigma_0 = 1.5$. We obtain $\sigma^2_{mj}$ (K19) from $\sigma^2_{b,1}$ (K17), and the latter from $\sigma^2_{bc}$ and $\sigma^2_{c,1}$, using (C1). Then, for $b = 0.41$, $c = 0.75$ and trimming fraction $p = 0.5$:

$$
\begin{aligned}
\sigma^2_{bc} &= \sigma^2_j\Big[1 - \frac{\Phi^{-1}(c)\phi\big(\Phi^{-1}(c)\big) - \Phi^{-1}(b)\phi\big(\Phi^{-1}(b)\big)}{c-p} - \\
&\Big(\frac{\phi\big(\Phi^{-1}(c)\big) - \phi\big(\Phi^{-1}(b)\big)}{c-p}\Big)^2\Big] \\
&= 1.5^2\Big[1 - \frac{\Phi^{-1}(0.75)\phi\big(\Phi^{-1}(0.75)\big) - \Phi^{-1}0.41\phi\big(\Phi^{-1}0.41\big)}{0.75-p} - \\
&\Big(\frac{\phi\big(\Phi^{-1}(0.75)\big) - \phi\big(\Phi^{-1}0.41\big)}{0.75-0.5}\Big)^2\Big] \\
&= 0.15,
\end{aligned} \tag{L7}
$$

and, analogously, for $c = 0.75$, $\sigma^2_{c,1} = \sigma^2_{0.75,1} = 0.54$.

To obtain $\sigma^2_{b,1}$, we first define truncated means, $\mu_{bc}$ and $\mu_{c,1}$, using (D6), with $Q_{bc}$ already defined in L6), giving

$$
\mu_{bc} = -\sigma_0 Q_{bc} = -1.5 \times 0.21 = 0.31, \tag{L8}
$$

and, in the same manner, $\mu_{c,1} = -\sigma_0 Q_{c,1} = -1.5 \times -1.27 = 1.91$.

Filling in (K17), with fractions $f_1 = (f_{0.5,c}/f_{0.5,1})$ and $f_1 = (f_{c,1}/f_{0.5,1})$, we obtain

$$
\begin{aligned}
\sigma^2_{b,1} &= f_1\sigma^2_{bc} + f_2\sigma^2_{c,1} + f_1 f_2(\mu_{bc} - \mu_{c,1})^2 \\
&= 0.5 \times 0.15 + 0.5 \times 0.54 + 0.25(0.31 - 1.91)^2 \\
&= 0.98.
\end{aligned} \tag{L9}
$$

To obtain the mirrored placebo distribution mean, $\sigma^2_{m0}$, we first define $\mu_{bd}$ as the sum of weighted fraction means, with $\mu_{b,1} = f1\mu_{bc} + f2\mu_{c,1} = 0.5 \times 0.31 + 0.5 \times 1.91 = 1.11$, and the mirrored mean, $\mu'_{b,1}$, with $\mu'_{b,1} = -\mu_{b,1} + 2\sigma_0 Z_b = -1.11 + 2 \times -0.23 = -1.80$, where $Z_b = -0.23$ is the normal quantile corresponding to $b = 0.41$ and also the mean of the mirrored distribution, $\mu_{m0}$.

Filling in (K19), we obtain:

$$
\begin{aligned}
\sigma^2_{mj} &= \sigma^2_{b,1} + 0.25(\mu_{b,1} - \mu'_{b,1})^2 \\
&= 0.98 + 0.25(1.11 - -1.80)^2 \\
&= 3.10,
\end{aligned} \tag{L10}
$$

with $\sigma_{mj} = \sqrt{3.10} = 1.76$. We now fill in (L6) and obtain the adjusted estimator bias under placebo group rescaling and placebo group strong MNAR violation:

$$
\begin{aligned}
B_{at0} &= -\sigma_1 Q_{bd} - \Big( -\frac{\sigma_1\sigma_0}{\sigma_{m0}}(f_1 Q_{bc} + f_2 Q_{c,1}) - \frac{\sigma_1\sigma_0}{\sigma_{m0}} Z_b + \sigma_0 Z_b \Big) \\
&= -1 \times -0.8 - \Big( -\frac{1 \times 1.5}{1.76}(0.5 \times -0.21 + 0.5 \times -1.27) - \\
&\frac{1 \times 1.5}{1.76} - 0.23 + 1.5 \times -0.23 \Big) \\
&= 0.32.
\end{aligned} \tag{L11}
$$

### M Trade-off of location shift bias and strong MNAR bias under different trimming proportions

Let us once more consider the unadjusted TM estimator bias components, under dropout in the placebo group. From (B5) we have the location shift assumption bias:

$$
B_{tLS} = (\sigma_1 - \sigma_0)\frac{\phi\big(\Phi^{-1}(p)\big)}{1-p}, \tag{M1}
$$

and from (E4) the strong MNAR assumption bias:

$$
B_{tSM0} = -\frac{f_{0.5,c(0)}}{f_{0.5,1}}\sigma_0(Q_{0.5,c(0)} - Q_{bc(0)}), \tag{M2}
$$

with $b_0 = f_{0,c(0)} - f_{bc(0)}$, and

$$
f_{bc(0)} = \frac{f_{0,c(0)} f_{bc(0)}}{(f_{0,c(0)} - p_{d0})}. \tag{M3}
$$

In the main article, we primarily consider fixed 50% trimming. From (M1) and (M2, M3) we observe that $B_{tLS}$ and $B_{tSM}$, respectively, are also a function of the specified trimming proportion, $p$. More specifically, a larger $p$ will increase $B_{tLS}$, but decrease $B_{tSM}$. This bias trade-off is illustrated in Supplementary Figure S3, which shows the two bias components against the trimming fraction, $p$, for a scenario with 20% placebo group dropout ($p_d = 0.2$), spread across 75% of the distribution ($f_{ac} = 0.75$).

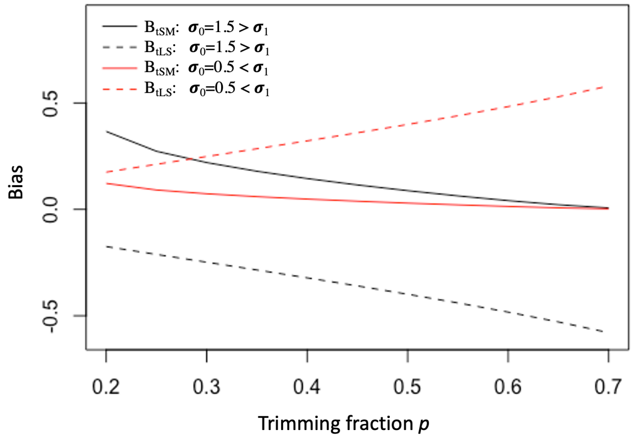

**Figure S3:** Location shift assumption bias $B_{tLS}$ (dashed line)and strong MNAR assumption bias $B_{tSM}$ (solid) across different trimming proportions $p$, under 20% placebo group dropout spread across 75% of the distribution ($f_{ac} = 0.75$), for placebo group SDs $\sigma_0 = 0.5$ (red) and $\sigma_0 = 0.5$ (black) and treatment group SD $\sigma_1 = 1$

#### N R code for obtaining unadjusted and unadjusted TM estimates

*TM.sim.func* is a function for simulating data from an RCT and performing an unadjusted and adjusted TM analysis under placebo group dropout and for 50% trimming. The function takes the following arguments.

- placebo group dropout proportion (*plD*)
- treatment group SD (*TRSD*)
- placebo group SD (*PLSD*)
- treatment effect size (*effTR*)
- sample size ($n$)
- dropout spread (e.g., *LOW* restricts all dropout to the lowest *plD*×100%)
- optional permutation function (when specifying *perm="YES"*, to obtain P value and 95% CI for unadjusted TM estimator

5mm
The function returns the following output.

- Specified treatment effect size (*effTR*)
- CCA estimate (*beta_c*)
- Unadjusted TM estimate (*beta_t*)
- Adjusted TM estimate, when rescaling the treatment group (*beta_t_resc_TR*)
- Adjusted TM estimate, when rescaling the placebo group (*beta_t_resc_PL*)
- The $P$-value and 95% confidence interval for the unadjusted TM estimate (*out_extra*)

```
TM.sim.func <- function(plD=0.2, TRSD=1, PLSD=1, n=10000,
                        effTR=0.5, opt="LOW", perm=FALSE,
                        a0 = 0, seed = NULL){
  if (!is.null(seed)) set.seed(seed)

  # patients randomized to treatment
  Tr <- rbinom(n,1,0.5)
  Tr <- sort(Tr, decreasing=FALSE)

  # generating normally distributed outcomes in treatment group,
  #with treatment effect effTR
  Yfin.mean.TR <- a0 + effTR
  Yfin.TR <- rnorm(length(which(Tr==1)),Yfin.mean.TR, TRSD)

  # generating normally distributed outcomes in placebo group
  Yfin.mean.PLAC <- a0
  Yfin.PLAC <- rnorm(length(which(Tr==0)), Yfin.mean.PLAC, PLSD)

  # joint outcome vector
  Yfin.TR <- sort(Yfin.TR)
  Yfin.PLAC <- sort(Yfin.PLAC)
  Yfin <- vector("numeric", n)
  Yfin[which(Tr==1)] <- Yfin.TR
  Yfin[which(Tr==0)] <- Yfin.PLAC

  # Number of dropouts (with plD specifying the dropout proportion)
  s.set <- round(plD*length(which(Tr==0)),0)

  # Generating dropout for four scenarios
```

```
  # Strict lower value dropout
if(opt=="LOW"){
  pl.out <- (seq(1,plD*length(which(Tr==0)),1))
  }
  # Dropout in bottom 50% of the distribution
if(opt=="LOWSCAT"){
  pl.out <- sample(seq(1,length(which(Tr==0))/2),s.set)
  }
  # Dropout in bottom 75% of the distribution
if(opt=="SCAT"){
  pl.out <- sample(seq(1,round(length(which(Tr==0))*0.75),0),s.set)}

  # Dropout across 100% of the distribution
if(opt=="ALL"){
  pl.out <- sample(seq(1,round(length(which(Tr==0))*0.75),0),s.set)}

# Generating dropout indicator
C.outs <- rep(1,n)
C.outs[c(pl.out)] <- 0

# Generating treatment group data with a column for outcome when
# fully observed (Yfin) and outcome after dropout (V4), where
# dropout are assigned a value (-Inf) lower than the smallest
# observed outcome. In this function there is no option for
# specifying treatment group dropout, so Yfin=V4 for the
# treatment group
dat.pT <- as.data.frame(cbind(IDs=seq(1,length(which(Tr==1)),1),
                              Yfin=Yfin[which(Tr==1)],
                              Tr=c(rep(1,length(which(Tr==1)))),
                              outs=C.outs[which(Tr==1)], V4=NA))
dat.pT$V4[which(dat.pT$outs==1)] <- dat.pT$Yfin[which(dat.pT$outs==1)]
dat.pT$V4[which(dat.pT$outs==0)] <- -Inf

# Generating placebo group data with a column for outcome when
# fully observed (Yfin) and outcome after dropout (V4), where
# dropouts are assigned a value (-Inf) lower than the smallest
# observed outcome.
dat.pP <- as.data.frame(cbind(IDs=seq(length(which(Tr==1))+1,
                                      length(Tr),1),
                              Yfin=Yfin[which(Tr==0)],
                              Tr=c(rep(0,length(which(Tr==0)))),
                              outs=C.outs[which(Tr==0)], V4=NA))

dat.pP$V4[which(dat.pP$outs==1)] <- dat.pP$Yfin[which(dat.pP$outs==1)]
dat.pP$V4[which(dat.pP$outs==0)] <- -Inf
```

```
# sorting outcomes per treatment group and 50% trimming
dat.ex.tr.sort <- dat.pT[order(dat.pT$V4),]
dat.ex.plac.sort <- dat.pP[order(dat.pP$V4),]

trim.frac.tr <- round(0.5*nrow(dat.ex.tr.sort),0)
trim.frac.plac <- round(0.5*nrow(dat.ex.plac.sort),0)

dat.ex.tr.trim <- dat.ex.tr.sort[-(1:trim.frac.tr),]
dat.ex.plac.trim <- dat.ex.plac.sort[-(1:trim.frac.plac),]

# CCA estimate
compdat <- rbind(dat.pT, dat.pP)
compdat <- compdat[which(compdat$outs==1),]
beta_c <- summary(lm(Yfin~Tr, compdat))$coefficients[2,1]

# Unadjusted TM estimate
beta_t <- mean(dat.ex.tr.trim$Yfin)-mean(dat.ex.plac.trim$Yfin)

# Adjusted TM estimate, obtained by rescaling the treatment group
  # extrapolating full sample SD from placebo group trimmed fraction
sd.tr.PL <- sqrt((sd(dat.ex.plac.trim$Yfin)^2)/(1-2/pi))
  # unscaled outcome
x1 <- dat.ex.tr.trim$Yfin
  # mirrored distribution of unscaled outcome
x2 <- min(x1) - (abs((min(x1)-x1)))
  # pooled distribution, after mirroring
x3a <- c(x1,x2)
  # rescaling mirrored treatment group distribution
x3 <- x3a - mean(x3a)
x4 <- x3/(sd(x3)/sd.tr.PL)
x4 <- x4 - mean(x4)
x5 <- x4 + mean(x3a)
  # obtaining original, now rescaled values
x6 <- x5[1:length(x1)]

  # replacing unscaled values in dataframe with rescaled ones
dat.ex.tr.trim2 <- dat.ex.tr.trim
dat.ex.tr.trim2$Yfin <- x6

beta_t_resc_TR <- mean(dat.ex.tr.trim2$Yfin)-
                  mean(dat.ex.plac.trim$Yfin)

# Adjusted TM estimate, obtained by rescaling the placebo group,
```

```
# using the SD of the fully observed treatment group
sd.tr.TR <- sd(dat.ex.tr.sort$Yfin)
# unscaled outcome
x1 <- dat.ex.plac.trim$Yfin
# mirrored distribution of unscaled outcome
x2 <- min(x1) - (abs((min(x1)-x1)))
# pooled distribution, after mirroring
x3a <- c(x1,x2)
# rescaling mirrored treatment group distribution
x3 <- x3a - mean(x3a)
x4 <- x3/(sd(x3)/sd.tr.TR)
x4 <- x4 - mean(x4)
x5 <- x4 + mean(x3a)
# obtaining original, now rescaled values
x6 <- x5[1:length(x1)]
# replacing unscaled values in dataframe with rescaled ones
dat.ex.plac.trim2 <- dat.ex.plac.trim
dat.ex.plac.trim2$Yfin <- x6

beta_t_resc_PL <- mean(dat.ex.tr.trim$Yfin)-
                    mean(dat.ex.plac.trim2$Yfin)

# output with true treatment effect, CCA estimate, TM estimate,
# adjusted TM estimate under treatment group rescaling,
# adjusted TM estimate under placebo group rescaling
out <- c(effTR, beta_c, beta_t, beta_t_resc_TR, beta_t_resc_PL)
out.extra <- c(0,0,0)

# permutation function for confidence intervals and P-value
# unadjusted TM estimator
perm.ff <- function(dat.trim){
  new.tr <- sample(dat.trim$Tr, replace=FALSE)
  perm.est <- summary(lm(dat.trim$Yfin~new.tr))$coefficients[2,1]
  return(perm.est)
}

if(perm==TRUE){
  perm.testing <- replicate(1000, perm.ff(dat.trim))
    # obtaining P value
  Pval <- (length(perm.testing)-sum(beta_t>perm.testing))/
                length(perm.testing)
    # obtaining confidence interval
  sd.perm <- sd(perm.testing)
  conf.int <- c(beta_t-sd.perm*1.96, beta_t+sd.perm*1.96)
  out.extra <- c(Pval, conf.int)
}
```

```
  # output with true treatment effect, CCA estimate, TM estimate,
  # Adjusted TM estimate under treatment group rescaling, Adjusted
  # TM estimate under placebo group rescaling, TM estimate P value,
  # and TM estimate 95% CI
  if(perm==TRUE){
    out <- c(effTR, beta_c, beta_t, beta_t_resc_TR, beta_t_resc_PL,
             out.extra)
  }

  return(out)
}
```

### O Application using the CoBalT RCT: BDI-II score distributions across time and treatment groups

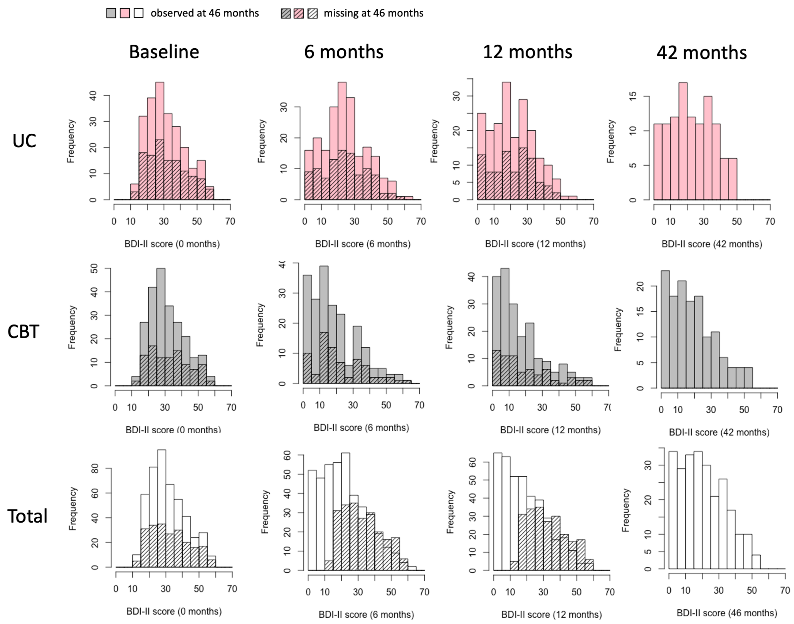

**Figure S4:** BDI-II score distributions at baseline, 6, 12 and 42 months for the usual care placebo group (UC, pink), the CBT treatment group (CBT, grey), and both groups jointly (Total, white). Outcome values missing at 42 months are indicated with a diagonal stripe pattern for each of the preceding time points.

### P Inferring the full sample SD from the SD observed under dropout, and calculating bias for the CoBalT application (R code)

For a given dropout scenario, the TM estimator bias can be quantified using the formulae detailed in Sections 2.3 and 2.4 of the main text (see also Appendices B-E). Both the locations shift assumption bias and strong MNAR bias are a function of the full sample SDs, which remain unobserved in the presence of dropout. Under the assumption of normally distributed outcomes these can, however, for a given dropout mechanism, be inferred from the observed SDs. The specified dropout mechanism has implications for the underlying full sample SDs. Consider, for example, the scenario in Figure 2.2 of the main text, where we have 41.9% fully directional dropout in the treatment (CBT) group and 52.3% entirely homogeneous dropout in the comparator (UC) group, which satisfies and violates the strong MNAR assumption in the treatment and comparator group, respectively. While the observed comparator group SD, under homogeneous dropout, is representative of the full sample SD, the observed treatment group SD, under assumption of highest value dropout, is the SD of a partial distribution. Under assumption of normality, we can consider this the SD of a normal distribution truncated at 41.9%, and infer the corresponding full sample SD.

Consider the variance of a truncated normal distribution, $\sigma^2_{ab}$,for a given fraction of the distribution, $f_{a,b}$ with left-truncation at $a$ and right-truncation at $b$ (see also (C1), Appendix C):

$$\begin{aligned}\sigma^2_{ab} = \sigma^2\Big[1 - &\frac{\Phi^{-1}(b)\phi\big(\Phi^{-1}(b)\big) - \Phi^{-1}(a)\phi\big(\Phi^{-1}(a)\big)}{b-a} - \\ &\Big(\frac{\phi\big(\Phi^{-1}(b)\big) - \phi\big(\Phi^{-1}(a)\big)}{b-a}\Big)^2\Big],\end{aligned} \tag{P1}$$

with corresponding truncated mean (see also (B2), Appendix B):

$$\mu_{ab} = \mu - \sigma\frac{\phi\big(\Phi^{-1}(b)\big) - \phi\big(\Phi^{-1}(a)\big)}{b-a}. \tag{P2}$$

Let $f_{0,c}$ denote the fraction of the outcome distribution unaffected by dropout, and $f_{c,1}$ the fraction of the distribution affected by dropout (note that the location of dropout, be it towards the top or bottom of distribution, does not impact the observed or full sample SD). The corresponding variances, $\sigma^2_{0,c}$ and $\sigma^2_{c,1}$, can be pooled to obtain the total variance. Note that the variance of the fraction affected by dropout, $\sigma^2_{c,1}$, under the assumption of homogeneous dropout in this fraction, is equal to the variance in absence of dropout.

Let $f_1 = \frac{f_{c,1}-p_d}{1-p_{d0}}$ and $f_2 = \frac{f_{0,c}}{1-p_{d0}}$ denote the proportions of observations, after dropout, in $f_{c,1}$, and $f_{0,c}$, respectively. The observed variance, after dropout, can be written as the pooled variance of the two fractions (see also (K16), Appendix K.5):

$$\sigma^2_o = f_1\sigma^2_{0,c} + f_2\sigma^2_{c,1} + f_1 f_2(\mu_{0,c} - \mu_{c,1})^2, \tag{P3}$$

Assuming, without loss of generality, a 0-centered distribution, and using (P1) and (P2), we write (P3) as a function of the full variance, $\sigma^2$:

$$
\begin{aligned}
\sigma_o^2 = f_1\sigma^2\Big[1 - \frac{\Phi^{-1}(c)\phi\big(\Phi^{-1}(c)\big)}{c} - \Big(\frac{\phi\big(\Phi^{-1}(c)\big)}{c}\Big)^2\Big] + \\
f_2\sigma^2\Big[1 + \frac{\Phi^{-1}(c)\phi\big(\Phi^{-1}(c)\big)}{1-c} - \Big(\frac{\phi\big(\Phi^{-1}(c)\big)}{1-c}\Big)^2\Big] + \\
f_1f_2(-\sigma\frac{\phi\big(\Phi^{-1}(c)\big)}{c} - \sigma\frac{\phi\big(\Phi^{-1}(c)\big)}{1-c})^2.
\end{aligned}
\tag{P4}
$$

By filling in the observed variance, specifying the dropout spread and proportion, we can solve for $\sigma$ and infer the unobserved, full sample SD. Below, R code is given for the implementation of this. Additionally, code is provided for functions that calculate the location shift assumption bias and strong MNAR assumption bias. These are used to calculate the expected bias for the TM estimate of the CBT treatment effect in the CoBalT data, for three different dropout mechanisms.

```
# function for calculating full sample SD from observed
# variance (obs.var), dropout spread (DS) and dropout
# proportion (dr), under assumption of normality
# implements equation (P4)
SD.func.extr <- function(DS, dr, obs.var){

  c <- DS
  fr1 <- (c-dr)/(1-dr)
  fr2 <- (1-c)/(1-dr)

  A <- (1- ((qnorm(c)*dnorm(qnorm(c)))/(c)) -
          ((dnorm(qnorm(c))/(c)))^2)
  B <- (1- ((-qnorm(c)*dnorm(qnorm(c)))/(1-c))-
          ((-dnorm(qnorm(c))/(1-c)))^2)
  C <- -(dnorm(qnorm(c)))/(c)
  D <- (dnorm(qnorm(c)))/(1-c)

  fsd <- function(sigf) {((fr1*A*sigf^2 + fr2*B*sigf^2 +
                          fr1*fr2*(C*sigf-D*sigf)^2))-
                            obs.var}

  fullSD <- uniroot(fsd, lower=0.1, upper=30)$root
  return(fullSD)}

# Function for calculating the location shift assumption bias
# for a given treatment group SD (TRSD) and comparator group SD
```

```
# (PLSD), for a specified trimming fraction (trim.frac)
# Implements equation (B5) in Appendix (B)
LSA.bias <- function(TRSD, PLSD, trim.frac){
  -(TRSD - PLSD) * (dnorm(qnorm(1-trim.frac))-dnorm(qnorm(0)))/
                        (1-trim.frac)}

# Function for calculating the strong MNAR assumption bias for a
# given treatment group (group="TR or group="PL), from the full
# sample SD (SD), the dropout spread (DSP), the dropout proportion
# (plD) and the trimming fraction (trimf).
# implements equations (E4) and (E5) in Appendix E
bias.drop <- function(SD=1,DSP=0.9, plD=0.2, trimf=0.55,
                group="TR"){

  DS <- 1-DSP
  calc.frac <- ((1-DS) * ((1-DS)-trimf) / ((1-DS)-plD))

  if (DSP <= trimf){
    calc.frac <- 0
  }
  calc.frac

  b.prop <- 1-trimf
  c.prop <- DS

  bst.prop <- DS+calc.frac
  a <- qnorm(DS+calc.frac,0,1)
  b <- qnorm(DS,0,1)
  a1 <- qnorm(1-trimf,0,1)

  F1 <- (((1-trimf)-DS)/(1-trimf))

  if(DSP<=trimf){
    a <- qnorm(1-trimf,0,1)
    b <- qnorm(0,0,1)
    a1 <- qnorm(1-trimf,0,1)
    F1 <- 1
  }

  elPL <- ((((dnorm(a1)-dnorm(b))/
                    (pnorm(a1)-pnorm(b)))*-SD)-
        ((dnorm(a)-dnorm(b))/
                (pnorm(a)-pnorm(b))*-SD)) *F1
  elTR <- (((dnorm(a)-dnorm(b))/
                (pnorm(a)-pnorm(b))*-SD) -
```

```
        ((((dnorm(a1)-dnorm(b))/
                (pnorm(a1)-pnorm(b)))*-SD))) *F1

  if(group=="TR"){
    el <- elTR
  } else { if (group=="PL"){
    el <- elPL}
  }

  if(group!="PL" & group!="TR"){
    el <- NA}

  el

  return(el)}

# Calculating expected bias for the CBT treatment effect
# estimate in CoBalT data, under 53.3% trimming, for three
# different dropout scenarios.

# comparator (UC) group with homogeneous dropout, treatment
# (CBT) group with completely directional dropout in
# upper 41.9%
  # comparator (UC) group SD
SD.PL.1 <- SD.func.extr(DS=0.999, dr=0.533,
            obs.var=13.2^2) # 13.12
  # treatment (CBT) group SD
SD.TR.Dir <- SD.func.extr(DS=0.419, dr=0.419,
            obs.var=13.8^2) # 21.54
  # Location shift assumption bias
LS_bias_a <- LSA.bias(SD.TR.Dir, SD.PL.1, 0.533) # -7.17
  # Strong MNAR assumption bias in comparator (UC) group
SM_bias_PL_a <- bias.drop(SD=SD.PL.semi_Hom, DSP=1, plD=0.533,
                          trimf=0.533, group="PL") #-10.36
  # Total bias
tot_bias_a <- LS_bias_a+SM_bias_PL_a # -17.53

# comparator (UC) group with homogeneous dropout, treatment
# (CBT) group with completely directional dropout in upper 60%
  # comparator (UC) group SD
SD.PL.1 <- SD.func.extr(DS=0.9999, dr=0.533,
                    obs.var=13.2^2)
  # treatment (CBT) group SD
SD.TR.semi_Dir1 <- SD.func.extr(DS=0.6, dr=0.419,
                          obs.var=13.8^2)
```

```
  # Location shift assumption bias
LS_bias_b <- LSA.bias(SD.TR.semi_Dir1, SD.PL.1, 0.533) # -1.14
  # Strong MNAR assumption bias in comparator (UC) group
SM_bias_PL_b <- bias.drop(SD=SD.PL.semi_Hom, DSP=1, plD=0.533,
                        trimf=0.533, group="PL") #-10.36
  # Strong MNAR assumption bias in treatment (CBT) group
SM_bias_TR_b <- bias.drop(SD=SD.TR.semi_Dir2, DSP=0.6,
                plD=0.419,trimf=0.533,group="TR") # 0.41
  # total bias
tot_bias_b <- LS_bias_b+SM_bias_PL_b+SM_bias_TR_b #-11.09

# comparator (UC) group with mostly homogeneous dropout in
# upper 80% of the distribution, and treatment (CBT) group with
# mostly directional dropout in upper 60%
  # comparator (UC) group SD
SD.PL.semi_Hom <- SD.func.extr(DS=0.8, dr=0.533,
                    obs.var=13.2^2)
  # treatment (CBT) group SD
SD.TR.semi_Dir2 <- SD.func.extr(DS=0.6, dr=0.419,
                    obs.var=13.8^2)
  # Location shift assumption bias
LS_bias_c <- LSA.bias(SD.TR.semi_Dir2, SD.PL.semi_Hom, 0.533) # -2.01
  # Strong MNAR assumption bias in comparator (UC) group
SM_bias_PL_c <- bias.drop(SD=SD.PL.semi_Hom, DSP=0.8, plD=0.533,
                        trimf=0.533,group="PL") #-5.50
  # Strong MNAR assumption bias in treatment (CBT) group
SM_bias_TR_c <- bias.drop(SD=SD.TR.semi_Dir2, DSP=0.6, plD=0.419,
                        trimf=0.533,group="TR") # 0.41
  # Total bias
tot_bias_c <- LS_bias_c+SM_bias_PL_c+SM_bias_TR_c #-7.10
```
